# Data-Driven Multiscale Analysis of the HIV Epidemic in the USA: Structural and Practical Identifiability Across Epidemiological Scales

**DOI:** 10.64898/2026.07.30.26359343

**Authors:** Leila Mirsaleh Kohan, Maia Martcheva, Necibe Tuncer

## Abstract

We present a multiscale model of HIV that couples within-host viral dynamics with population-level transmission to capture the interplay between individual infection and epidemic spread. The model is structured by treatment age, allowing viral load to influence both infectiousness and progression to AIDS. The model is fitted using both clinical data (viral load and target cell counts of treated individuals) and epidemiological data (HIV incidence, diagnoses, and AIDS classifications). We derive the basic reproduction number and establish threshold conditions for the existence and stability of disease-free and endemic equilibria. Structural identifiability is assessed via input-output equations, showing that the multiscale model is identifiable when the initial number of treated individuals is set to zero and the AIDS death rate is assumed known. Parameters are estimated sequentially: within-host parameters are obtained via nonlinear mixed-effects modeling of clinical data, followed by estimation of population-level parameters using CDC surveillance data. Numerical simulations are performed using a finite-difference scheme with Picard iteration, and practical identifiability is evaluated via Monte Carlo simulations across varying noise levels. Results indicate that current strategies are unlikely to meet the 2030 targets, while increasing diagnosis rates and reducing transmission from diagnosed individuals could significantly alter epidemic trajectories. These findings highlight the importance of multiscale modeling and identifiability in informing effective HIV intervention strategies.

## 1 Introduction

Human immunodeficiency virus (HIV) remains a major global public health challenge despite substantial advances in treatment and prevention. The development of effective antiretroviral therapies (ART) has transformed HIV from a fatal disease into a manageable chronic condition [1]. Nevertheless, achieving the ambitious goal of ending the HIV epidemic by 2030, as set by global initiatives such as UNAIDS and the United States Ending the HIV Epidemic (EHE) plan [2–6], remains a significant challenge due to persistent transmission [7], viral mutation and drug resistance [8], and barriers to sustained treatment and care [9, 10]. Mathematical models have played a crucial role in understanding HIV dynamics and informing intervention strategies [11]. Early models focused on either within-host viral dynamics [12] or between-host epidemiological transmission [13]. However, the interconnected nature of these processes necessitates a more integrated approach. Multiscale nested immuno-epidemiological models have emerged as a powerful tool to capture synergistic interactions between individual-level immune responses and population-level transmission dynamics [14]. Early multiscale models of HIV were developed by DebRoy and Martcheva [15], while more sophisticated frameworks subsequently considered factors such as HIV super-infection [16], coinfection [17], age-since-infection multiscale HIV dynamics [18], network immuno-epidemiological dynamics [19], and HIV-opioid and co-affection dynamics [20], together with drug resistance, viral evolution, and treatment in HIV-positive individuals, and their collective impact on epidemic pathways.

Multiscale immuno-epidemiological models of HIV were reviewed in [21] up to 2017, and they have continued to be a significant modeling tool since then [19, 22–26]. A critical issue in using multiscale HIV models is connecting them to data. Multiscale models can be linked to data by fitting to data sets from both scales. Unfortunately, this is rarely done [20], and multiscale models often have only theoretical significance. Parameter estimation in such models is an inverse problem and is often ill-posed, meaning that the same data can lead to multiple parameter sets that produce the same output [27].

Identifiability analysis assesses whether model parameters can be uniquely estimated from available data [28, 29]. We distinguish two major types of identifiability: structural identifiability and practical identifiability. Structural identifiability determines whether unique parameter estimates are theoretically possible under ideal noise-free conditions [28], while practical identifiability considers estimation uncertainty in the presence of data noise [29]. Despite its importance, identifiability analysis has received limited attention in multiscale HIV models, although public health decision-making relies on models with reliable parameter estimates [30].

While the tools for practical identifiability of nested multiscale models have been available for quite some time [31], the tools for structural identifiability remain in their early stages of development. Structural identifiability of ODE models is typically performed with multiple types of commercial software [32]. However, such software does not exist for partial differential equation models (PDE), including nested multi-scale models. The structural identifiability for PDE models has only recently begun to be investigated [27, 33], and is usually done analytically. Regarding nested immuno-epidemiological models, the only prior result on their structural identifiability that we are aware of is reference [34].

In this study, we develop and analyze a nested multiscale HIV model that couples within-host viral dynamics with population-level transmission through treatment-age structure. The model explicitly links viral load to both infectiousness and disease progression. We derive the basic reproduction number and establish threshold conditions for the existence and stability of disease-free and endemic equilibria. To ensure model reliability, we assess structural identifiability by deriving input–output equations and adapting differential algebra techniques to the PDE setting.

Parameter estimation is carried out sequentially: within-host dynamics are fitted to clinical data on viral load and CD4 cells using nonlinear mixed-effects modeling, followed by estimation of population-level parameters using CDC surveillance data on HIV incidence, diagnoses, and AIDS classifications. Practical identifiability is then evaluated through Monte Carlo simulations under varying noise levels. As an application, we use the fitted model to assess whether current intervention strategies are sufficient to achieve the 2030 EHE targets [4] and to identify potential pathways for improving epidemic control.

The remainder of this paper is organized as follows. Section 2 introduces the multiscale HIV model and the clinical and epidemiological data used for parameter estimation. In Section 3, we carry out structural identifiability analysis by deriving input–output equations and establishing conditions under which the model parameters are uniquely identifiable. Section 4 focuses on parameter estimation and practical identifiability, where we implement a sequential fitting approach combining nonlinear mixed-effects modeling of within-host dynamics with population-level data, and assess parameter uncertainty using Monte Carlo simulations. Section 5 presents the results of model fitting and validation. Section 6 introduces the concept of virtual epidemics, in which multiple epidemic trajectories are generated from data-consistent parameterizations of the multiscale model to assess uncertainty, explore dynamics, and evaluate the robustness of intervention strategies. Finally, Section 7 concludes the article with a discussion of the results and their implications for HIV epidemic control.

## 2 Multiscale HIV Model and Data

### 2.1 A Nested Multiscale HIV Model

We develop a multiscale HIV model that couples transmission dynamics at the population level with viral progression within individuals. The model extends standard compartmental approaches by incorporating treatment effects and the influence of viral load on both disease progression and transmission. The population is divided into four key compartments: susceptible individuals, *S*(*t*); infected individuals who are unaware of their status, *I*(*t*); diagnosed individuals who are being treated, *d*(*t, τ*) with treatment age *τ*; and individuals who have progressed to AIDS, *A*(*t*); all as functions of time *t*. In addition, the model captures the within-host viral dynamics of a treated individual through a system of differential equations for uninfected target cells, *T* (*τ*); infected cells, *T*_*i*_(*τ*); and viral load, *V* (*τ*). The system dynamics is governed by the following system of differential equations:

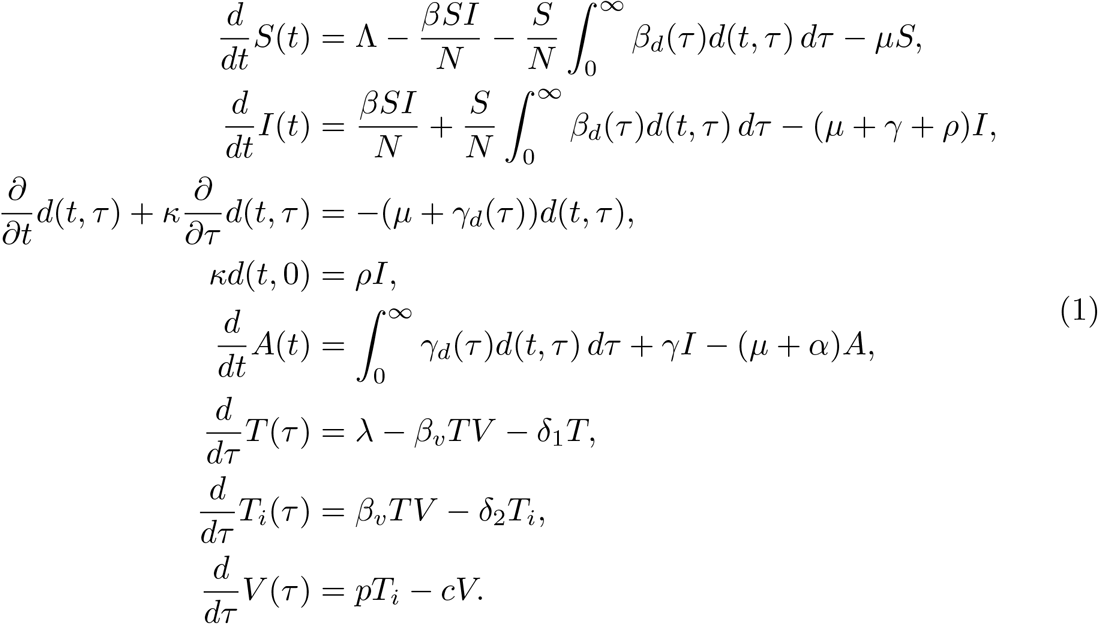

Because individuals in the AIDS class, *A*(*t*), are assumed to be too ill to contribute to new infections, they are excluded from the population contributing to disease incidence. Consequently, the total population size, *N*, is defined as *N* = *S*(*t*)+*I*(*t*) +*D*(*t*), where 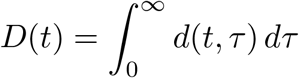 represents the total number of treated individuals at time *t*. The model incorporates the dependence of viral load in transmission and progression to AIDS rates, with 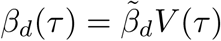 and 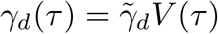.

The model captures key processes involved in HIV transmission and progression. Susceptible individuals *S*(*t*) are introduced into the sexually active population at a rate Λ. They are at risk of infection through contact with infected individuals *I*(*t*) who are unaware of their infectious status, modeled by the term 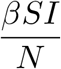, or through contact with treated individuals *d*(*t, τ*), represented by the integral term 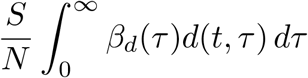. Natural death reduces the susceptible population at a rate *µS*(*t*). Infected individuals *I*(*t*) leave this compartment due to natural death *µI*(*t*), progression to AIDS *γI*(*t*), or diagnosis *ρI*(*t*). The treated individuals, represented by the density function *d*(*t, τ*), evolve with time *t* and treatment age *τ*. Epidemic time is measured in years and treatment age in weeks. Therefore, *κ* is the scaling factor between these two time scales. Individuals enter the treatment compartment through a boundary condition at a rate *ρI*(*t*). Treated individuals leave the compartment either due to natural death at rate *µd*(*t, τ*) or by progression to AIDS at rate *γ*_*d*_(*τ*)*d*(*t, τ*) which is determined by their viral load. The AIDS compartment *A*(*t*) includes individuals progressing from treatment failure at rate 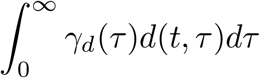 or directly from the infected class at rate *γI*(*t*). Individuals in this compartment experience an increase in mortality due to AIDS at rate (*α* + *µ*)*A*(*t*).

The within-host viral dynamics is captured by equations for uninfected target cells *T* (*τ*), infected target cells *T*_*i*_(*τ*), and viral load *V* (*τ*). These equations describe how viral particles interact with target cells: uninfected cells decrease due to infection at rate *β*_*v*_*TV*, infected cells increase due to infection but decrease due to clearance at rate *δ*_2_*T*_*i*_, and viral particles are produced by infected cells at rate *pT*_*i*_, while cleared at rate *cV*. The basic reproduction number, ℛ_*w*_, of the within-host model represents the average number of new infected cells produced by one infected cell in a population of uninfected target cells during the infected cell lifespan as infectious. It is given by:

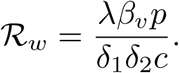

If ℛ_*w*_ *<* 1, the infection-free equilibrium is globally stable, leading to the eventual elimination of the infection [35]. if ℛ_*w*_ *>* 1, the infection-free state becomes unstable, resulting in persistent and chronic infection. The equilibrium values are the following.

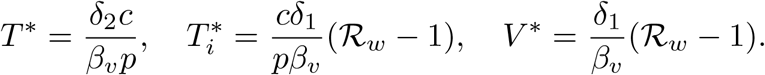

The multiscale model exhibits two equilibrium states: the disease-free equilibrium (DFE), where the disease is eliminated, and the endemic equilibrium (EE), where the disease persists at a constant level in the population. The detailed analysis of the multiscale model (1) is presented in Appendix A. The stability of the DFE is determined by the basic reproduction number, ℛ_0_, defined as

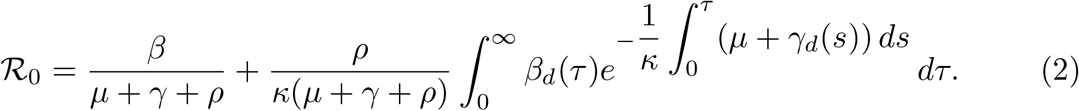

This basic reproduction number consists of three components; ℛ_0_ = ℛ_1_ +ℛ_2_ · ℛ_3_. The first term, 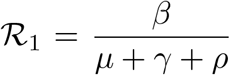, represents the average number of new infections caused by one infected individual before they are diagnosed or progress to AIDS. The second term, 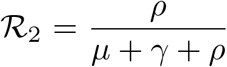, represents the probability that an infected individual is diagnosed. The integral 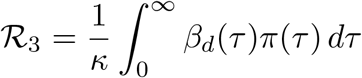 calculates the expected number of new infections caused by one individual after being diagnosed, over all possible durations of treatment. The DFE is locally stable if ℛ_0_ *<* 1. However, DFE becomes unstable if ℛ_0_ *>* 1, implying the potential for the disease to establish itself in the population. Furthermore, for the endemic equilibrium (EE) to be locally asymptotically stable, ℛ_0_ must not only be greater than 1, but also satisfy the condition 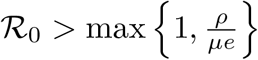 where *ρ* is the diagnosis rate and *µ* is the natural mortality rate. The quantity 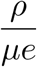 represents the balance between diagnosis efforts and the natural turnover of the population, with *e* arising from the characteristic equation analysis. The condition indicates that disease transmission must remain sufficiently strong to sustain endemicity despite ongoing diagnosis efforts and natural population turnover. Larger values of *ρ* increase this threshold, reflecting the stabilizing effect of more rapid diagnosis and treatment on disease spread. The initial conditions for the model are defined as *S*(0) = *S*_0_, *I*(0) = *I*_0_, *d*(0, *τ*) = *d*_0_(*τ*), *A*(0) = *A*_0_, *T* (0) = *T*_0_, *T*_*i*_(0) = *T*_*i*0_, and *V* (0) = *V*_0_. The definitions of all state variables and parameters of the model are provided in Table A1 and Table A2, respectively.

### 2.2 Multiscale Data

We estimate parameters of the multiscale model (1) using data at both population and individual scales. At the population scale, we use epidemic data reported by the CDC [36] and at the individual scale, we use HIV infection data reported in the Stanford HIV drug resistance database [37].

#### Population Scale Data

CDC reports estimated HIV incidence in the United States beginning in 2010 [36]. We obtained the HIV estimates for the period starting in 1981 by digitizing Figure 1 in [38] using the Grabit tool in MATLAB. The Figure 1 in [38] illustrates the changes in HIV incidence in the United States over nearly four decades. Data presented in Table B3 show that new HIV infections increased from around 20,800 cases in 1981 to a peak of 131,400 in 1984–1985. After that, the incidence declined to about 85,200 cases by 1990. Between 1991 and 2007, the incidence remained relatively steady, fluctuating between 49,500 and 58,600 cases per year. In recent years, the incidence decreased, reaching 36,200 new infections in 2019, a drop of about 73% from the peak in the mid-1980s. Table B4 shows the annual number of AIDS cases in the United States from 1985 to 2022 obtained from [36]. Initially, the cases increased sharply from 11,810 in 1985 to a peak of 73,970 in 1993. After introducing antiretroviral therapy in 1996, cases began to decline, reaching 56,500 in 1996. Table B5 shows HIV diagnosis data in the US from 2008 to 2019 as reported by CDC [36]. Over time, the number of diagnoses gradually declined from about 46,700 in 2008 to 36,350 in 2019.

**Fig. 1.**
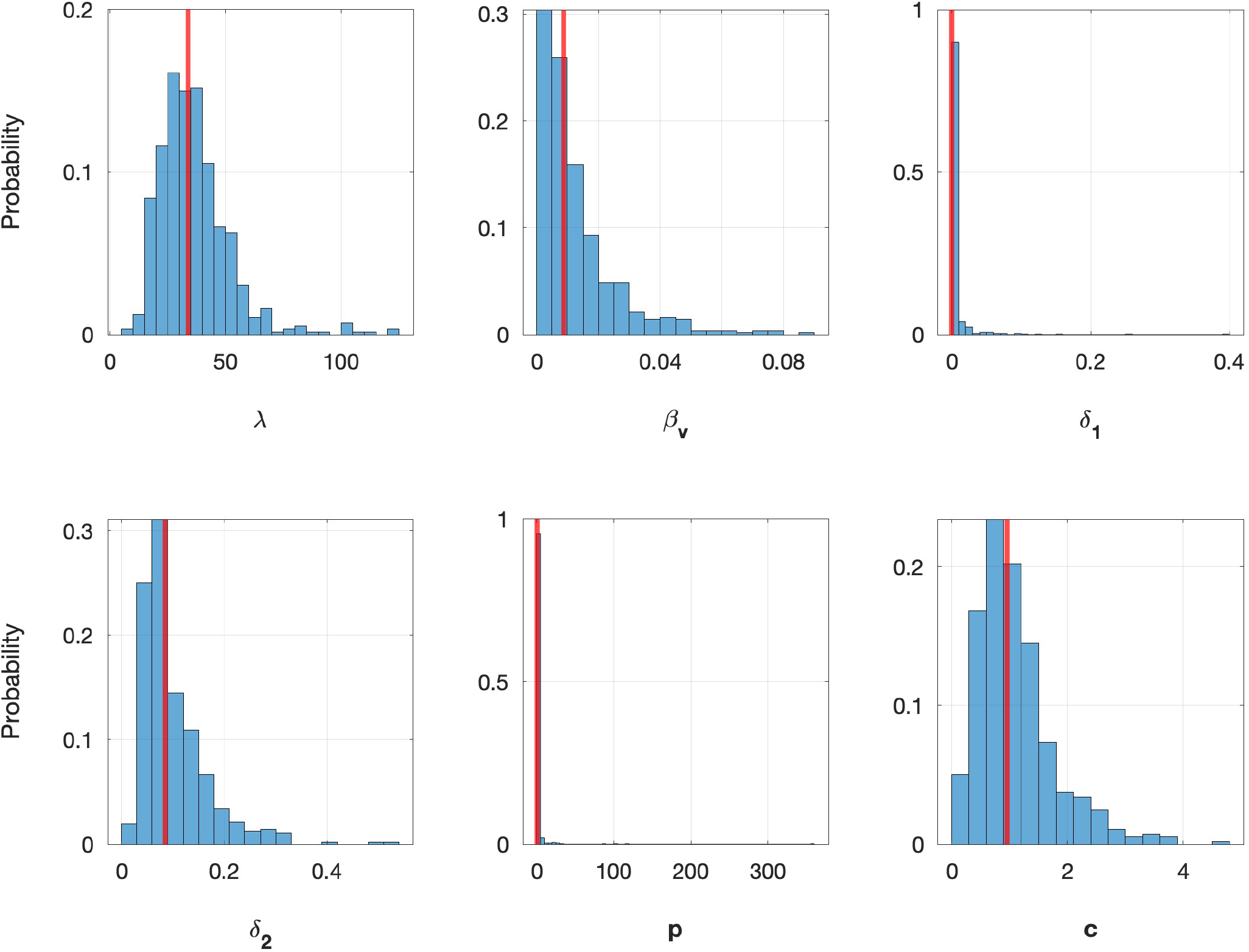
Probability distribution of within-host model parameters estimated by nonlinear mixed-effect modeling in Monolix; red vertical lines indicate the population means.

#### Individual Scale Data

To estimate the parameters of the within-host model, we utilize data from the Stanford HIV Drug Resistance Database, specifically the AIDS Clinical Trials Group (ACTG) 5257 study [39]. All participants had HIV-1 RNA levels greater than 1,000 copies/mL and the median CD4^+^ count was 308 cells/mm^3^ (interquartile range, 1170–425 cells/mm^3^). Participants were assigned to one of three regimens, each containing the nucleoside reverse transcriptase inhibitors (NRTIs) emtricitabine (200 mg/day) and tenofovir disoproxil fumarate (300 mg/day), combined with either atazanavir/ritonavir (a protease inhibitor regimen), darunavir/ritonavir (another protease inhibitor regimen), or raltegravir (an integrase inhibitor regimen). Virologic failure was defined as HIV-1 RNA levels that exceeded 1,000 copies/mL between weeks 16–24 or exceeded 200 copies/mL after week 24; among those who received raltegravir, 3% experienced virologic failure due to drug resistance. We selected 80 HIV patients from the study who had received NRTIs and PIs (protease inhibitors) [37]. The eight graphs in Figure B1 represent actual data sets from these 80 patients undergoing treatment, sourced from the Stanford HIV Drug Resistance Database. These graphs illustrate trends in CD4^+^ count (blue line) and viral load (red line) over time. The treatment phase, highlighted in gray, includes the following antiretroviral drugs: NRTIs (FTC, TDF) shown in blue, and PIs (ATV, RTV, DRV) shown in purple. The duration of data collection varies: some patients were followed for nearly 200 weeks, while others were followed for only 96 weeks, as shown in the Figure B1.

## 3 Structural Identifiability Analysis of Multiscale HIV Model

Before estimating the multiscale model parameters, we perform structural identifiability analysis of the HIV model (1). Structural identifiability determines whether unique parameter estimates can be obtained under the assumption of unlimited, noise-free data. For this work, we adopt an approach where we eliminate unobservable state variables and rewrite the system in terms of observed state variables and parameters.

This yields input-output equations expressed as differential-algebraic polynomials and differential-integral equations, where the outputs are related to the model parameters. The goal is to identify the model parameters, 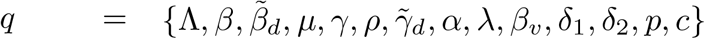, from observations of HIV incidence, AIDS classification, HIV diagnosis, total target cells, and viral load [36, 38, 39]. These observations are expressed in terms of the model variables as follows:

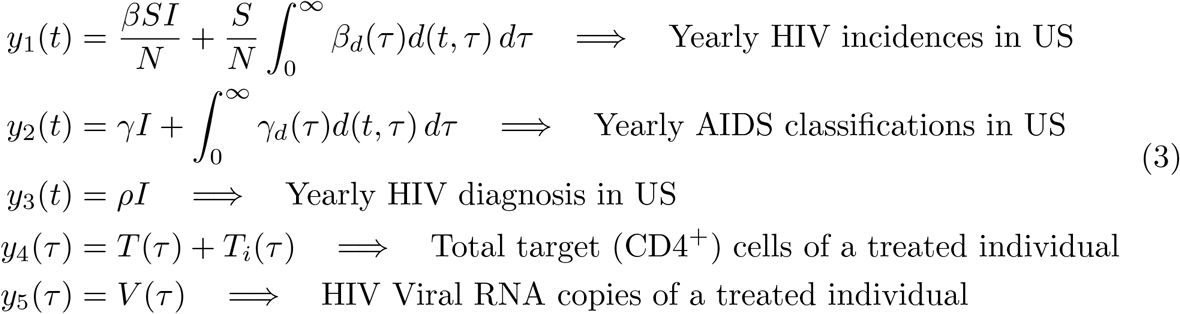

Let *x*(*t, τ*) = (*S*(*t*), *I*(*t*), *d*(*t, τ*), *A*(*t*), *T* (*τ*), *T*_*i*_(*τ*), *V* (*τ*)) be the state variables of the multiscale HIV model (1), and *g*(*x*(*t, τ*), *q*) denote the mapping of the state variables *x*(*t, τ*) (also implicitly of model parameters *q*) to observations, defined by,

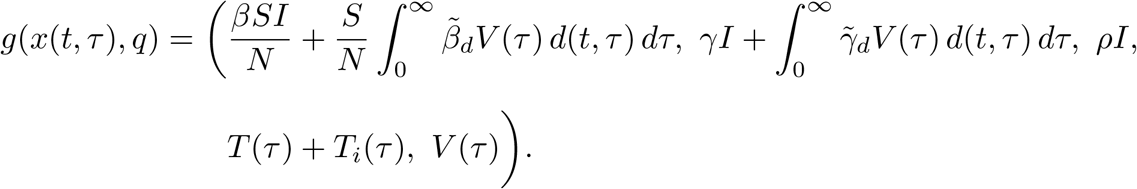

We begin by defining the structural identifiability for the multiscale model (1).

### Definition 1

Let *q* and 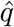 be distinct parameter vectors of the multiscale model (1). If

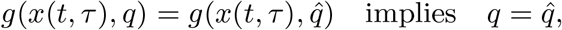

then the multiscale HIV model (1) is said to be structurally identifiable.

The model (1) is structurally identifiable if the mapping from the parameter space to the observations is one-to-one. To assess this, we derive the input-output equations. As a first step, we integrate the third equation of the HIV model (1) with respect to *τ*. Since *κ* lim_*τ*→∞_ *d*(*t, τ*) = 0 and *κd*(*t*, 0) = *ρI*(*t*), we obtain the following ordinary differential equation for *D*(*t*),

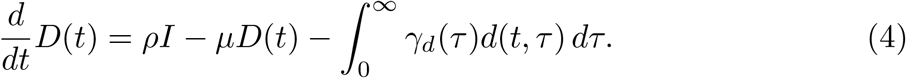

We reformulate the population-scale component of the multiscale model (1) in terms of observations *y*_1_(*t*), *y*_2_(*t*), *y*_3_(*t*).

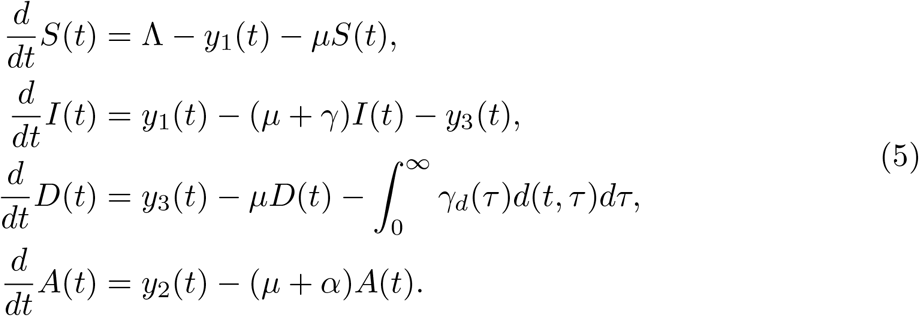

The total population *N* (*t*) = *S*(*t*) + *I*(*t*) + *D*(*t*) satisfies:

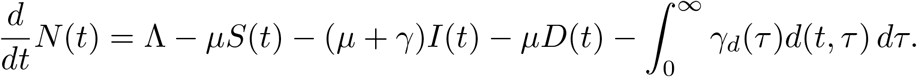

Rewriting this equation using the observations of AIDS classification, we get

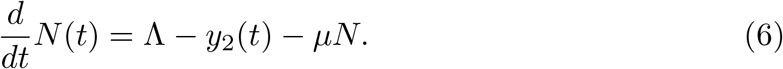

Multiplying the equation for *I*(*t*) by *ρ*, we obtain the following input-output equation.

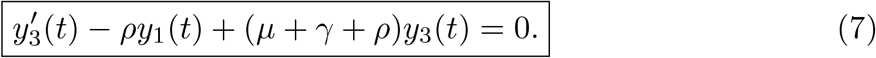

The observation, *y*_2_(*t*), of AIDS classifications can be rewritten as

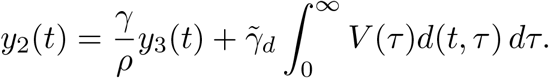

Isolating the integral term, we obtain:

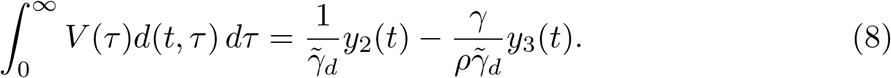

Then, the observation, *y*_1_(*t*), of the HIV incident can be written as

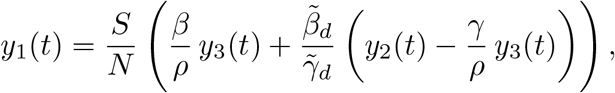

which can be further simplified to

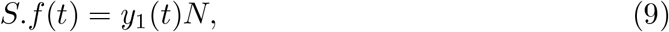

where 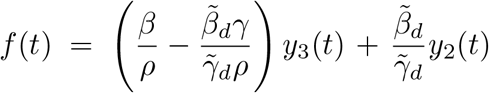. Differentiating both sides of (9) with respect to time, we get

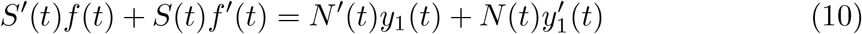

Substituting *S*^*′*^ = Λ − *y*_1_(*t*) − *µS*(*t*) and (6) into the above equation (3), we get,

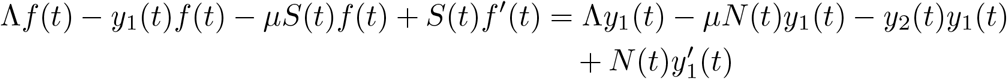

Now, using the original relation *S*(*t*)*f* (*t*) = *N* (*t*)*y*_1_(*t*), we obtain:

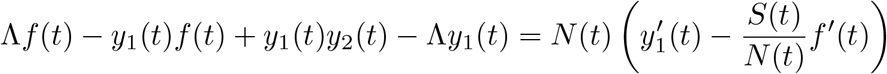

Now, using 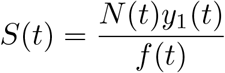 from the original identity, we substitute:

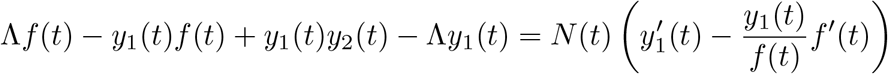

Solving for *N* (*t*), we obtain:

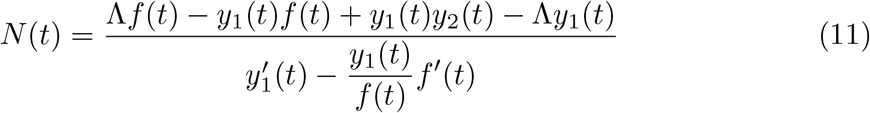

Finally, substituting (11) into the expression (6) for *N*^*′*^(*t*):

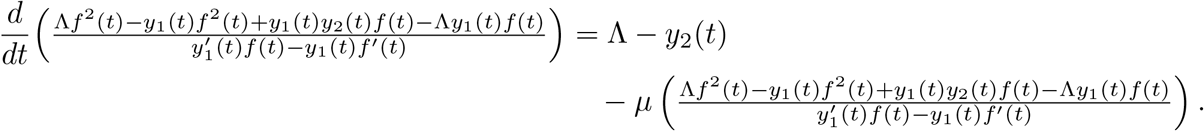

Differentiating the left-hand side yields the following expression, which we will use to derive the second input-output equation.

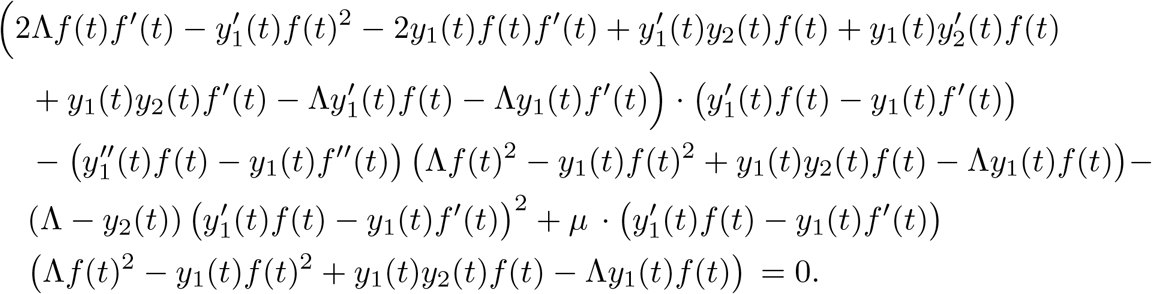

The following derivation is used to obtain the second input-output equation for the structural identifiability analysis of the multiscale HIV model. The resulting equation relates observable quantities to model parameters after eliminating unobservable state variables.

The final expression for the second input-output equation is obtained using *Mathematica* due to its algebraic complexity. The corresponding *Mathematica* expansion and the resulting full input-output equation are provided in Appendix A for reference.

Next, we derive the third input-output equation. We solve the partial differential equation (PDE) in (1) using the method of characteristics and obtain the following.

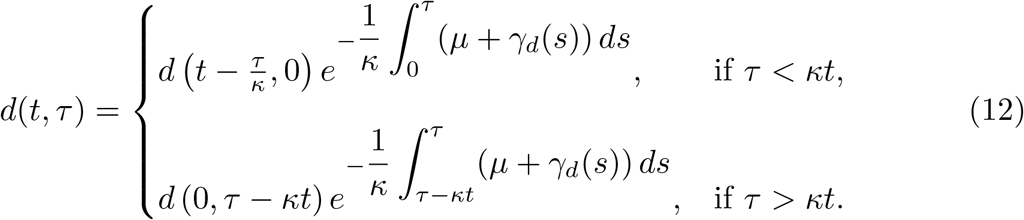

Substituting (12) into (8), we get

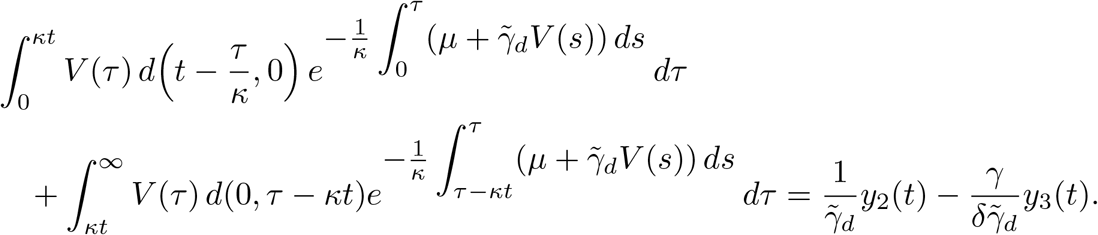

Since *d*(0, *τ*) = *d*_0_(*τ*), *d*(*t*, 0) = *y*_3_(*t*) and *V* (*τ*) = *y*_5_(*τ*) we obtain,

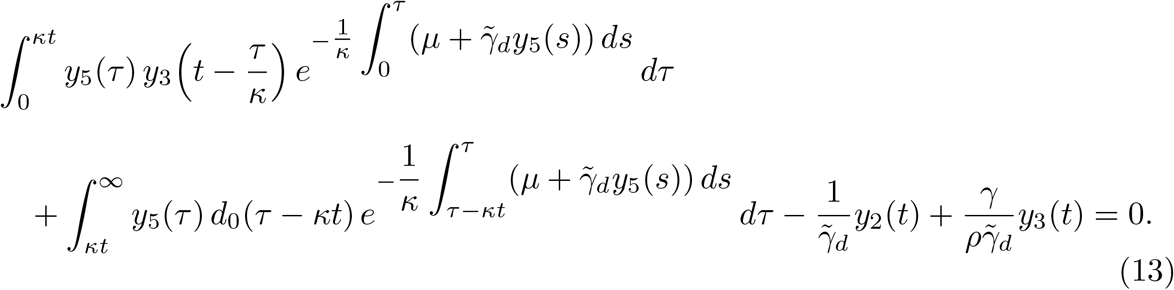

We simplify by defining

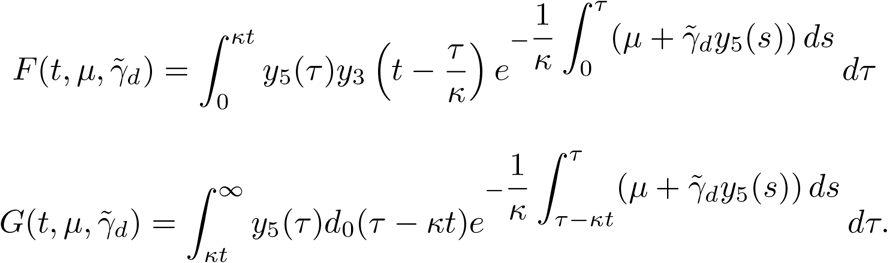

We then obtain the following input-output equation,

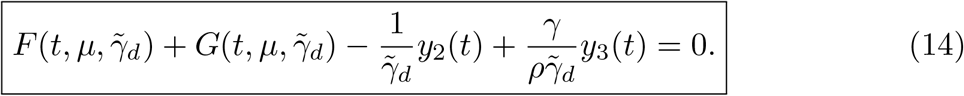

Next, we continue with the individual-scale component of the model (1). The observations at this scale are

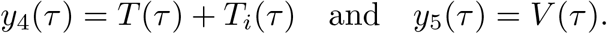

From the last equation of (1), we express *T*_*i*_(*τ*) in terms of *y*_5_(*τ*).

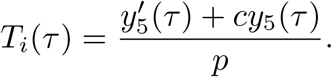

Adding the first and second equations of the individual scale component of the model (1), we obtain

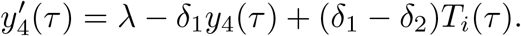

Substituting 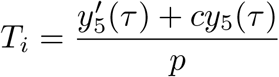, we obtain the fourth input-output equation,

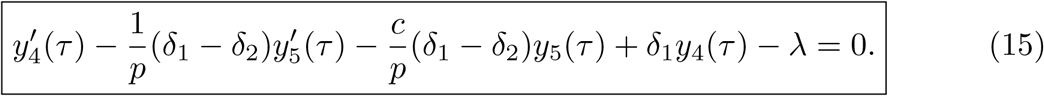

Isolating the infection term 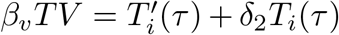, we get an expression for *T* (*t*) as

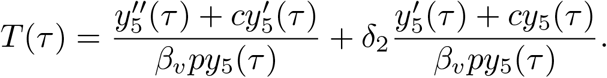

Thus, substituting *T* (*τ*) and *T*_*i*_(*τ*) into the expression for *y*_4_(*τ*), we get

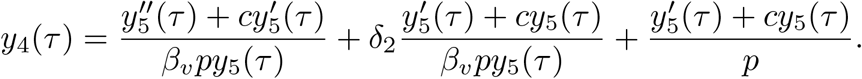

Multiplying both sides by *β*_*v*_*py*_5_(*τ*), we arrive at the final input-output equation.

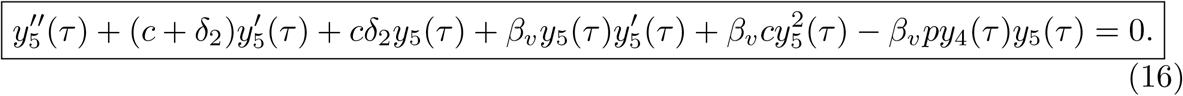

We proceed with the identifiability analysis by focusing on the within-host scale, represented by a system of ODEs. We derive input-output equations (15) and (16) in the form of differential polynomials. We then apply the following definition to determine the structural identifiability of the parameters [40].

### Definition 2

Let *q* and 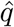 be any parameters of the individual-scale component of the multiscale model (1), and let *c*(*q*) represent the coefficients of the corresponding input-output equations (15),(16). The individual-scale component of the multiscale model (1) is structurally identifiable from the observations if and only if

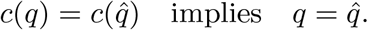

### Theorem 1

*The individual-scale component of the multiscale model* (1) *is structurally identifiable, from observations of HIV viral load and the total CD4 cell count*.

*Proof* Consider two sets of parameters, *q*^*′*^ = *{λ, β*_*v*_, *δ*_1_, *δ*_2_, *p, c}* and 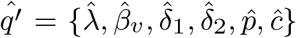, that yield the same observations, *y*_4_(*τ*) and *y*_5_(*τ*). Therefore, by setting, i.e., 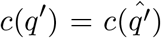, we obtain the following system of nonlinear equations.

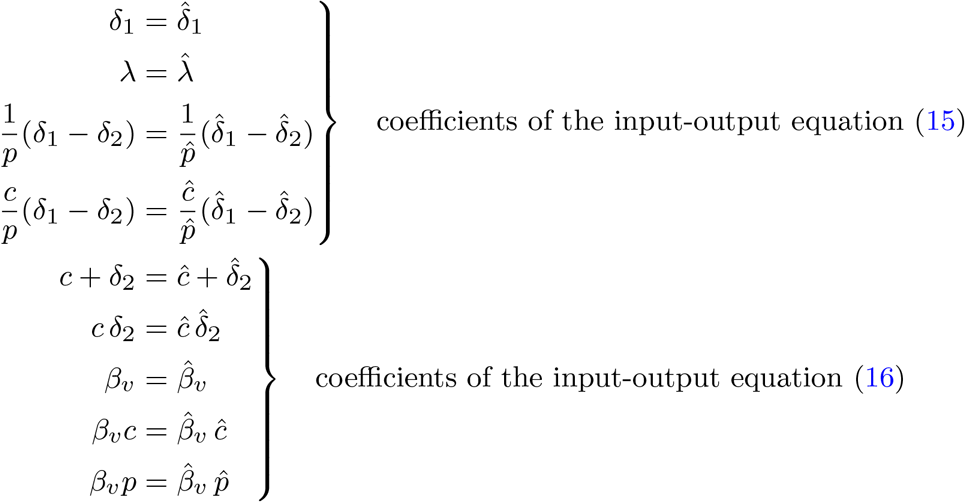

Parameters *λ, β*_*v*_ and *δ*_1_ are uniquely determined since 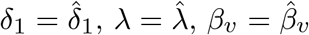. Further-more, since 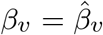, the equation 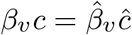, implies *c* = *ć*. Substituting into 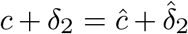 uniquely determines 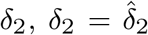. From 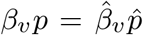, we obtain 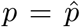. Thus, all parameters *{λ, β*_*v*_, *δ*_1_, *δ*_2_, *p, c}* are structurally identifiable. □

For the identifiability analysis of the multiscale model, we adopt a similar approach. It is important to note that not all the input-output equations of the multiscale system are differential polynomials; for instance, the input-output equation (14) is an integral-differential polynomial. Additionally, the parameter *α* is absent from all input-output equations of the multiscale system, namely (7), (C71), (14), (15), and (16). Therefore, it is clear that the parameter *α* is not identifiable from these observations, *y*_1_, *y*_2_, *y*_3_, *y*_4_, and *y*_5_. We perform the identifiability analysis by assuming that the initial number of treated individuals is zero, which is reasonable given that HAART treatment was not introduced until 1996 – 15 years after the HIV epidemic began in the USA.

### Theorem 2

*The multiscale model* (1) *is structurally identifiable from observations of HIV viral load, total CD4 count, yearly incidences, diagnoses, and AIDS classifications if the initial number of treated individuals is set to zero and the parameter α is assumed to be known*.

*Proof* Since *α* is given, let 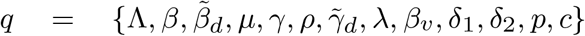 and 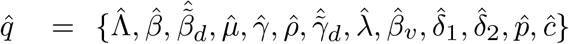 be two arbitrary parameter sets of the multiscale model. By Theorem 1, the within-host model is structurally identifiable; therefore, 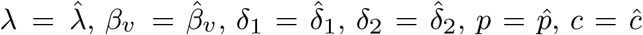. By equating coefficients of the input-output equations, specifically those expressed as differential polynomials, we obtain the following system of nonlinear equations. At the population scale, the input-output equations (7) and (C71) yield;

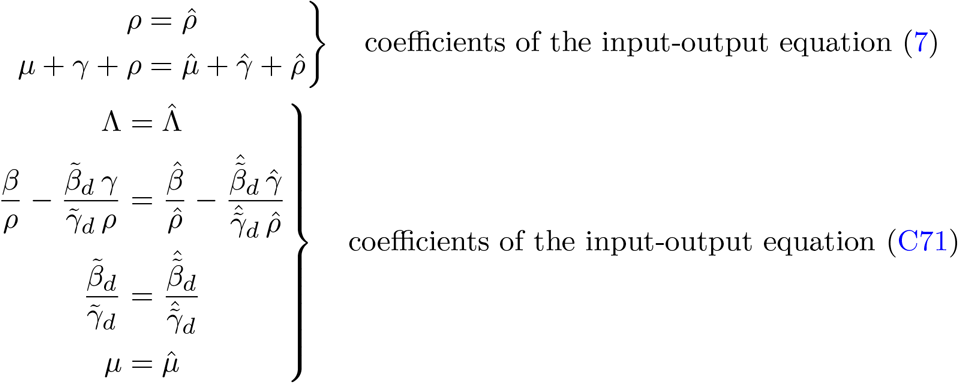

The parameters Λ, *µ* and *ρ* are determined uniquely, since 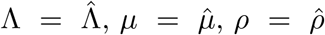. Furthermore, the equation 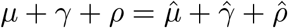 determines *γ* uniquely 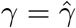.

Next, we are performing the identifiability analysis for the input-output equation, which is an integral-differential polynomial (14). Suppose that the two parameter sets *q* and 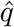 yield an identical input-output equation (14). Assuming the initial number of treated individuals is zero, that is, *d*(0, *τ*) = 0, the input-output equation (14) becomes

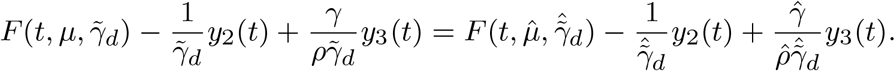

Note that, since 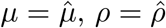 and 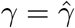, we have

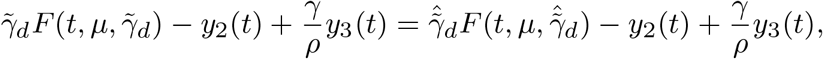

which is equivalent to

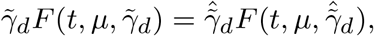

where

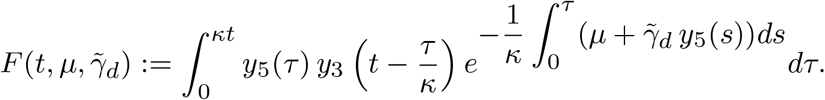

Assume for contradiction that 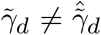, and the equality 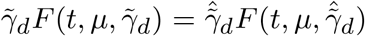 holds. By applying the Mean Value Theorem for integrals and using continuity of the functions involved, there exist *ξ*_1_, *ξ*_2_ ∈ (0, *κt*) such that

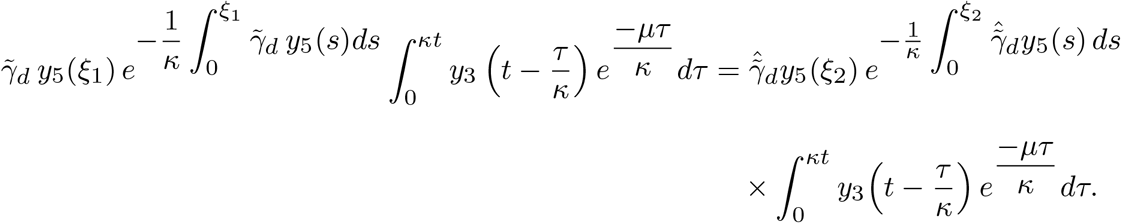

Since

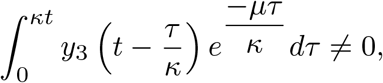

we cancel it from both sides to obtain

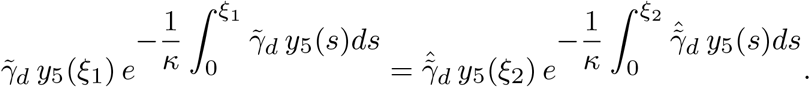

Now consider the function

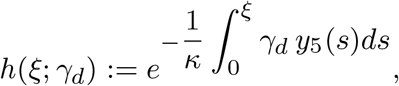

where *γ*_*d*_ is fixed. Differentiating with respect to *ξ*, we get

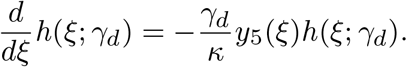

which implies

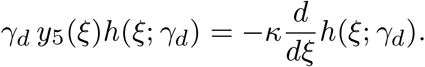

Applying this at *ξ*_1_ and *ξ*_2_ yields:

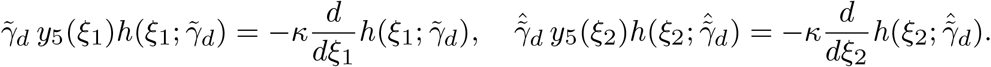

Hence, from the earlier equality, which is 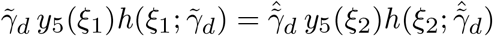

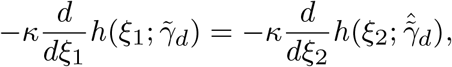

or

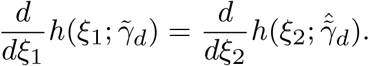

Integrating both sides over [0, *κt*] yields

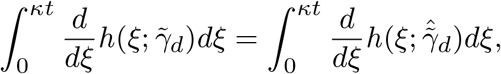

and by the Fundamental Theorem of Calculus,

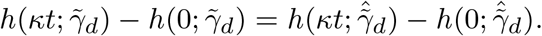

Since *h*(0; *γ*_*d*_) = 1 for any *γ*_*d*_, this implies

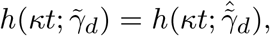

which implies

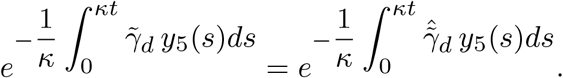

Taking logarithms yields

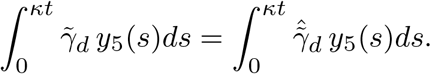

Since *y*_5_(*τ*) = *V* (*τ*) is positive, it follows that

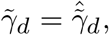

which contradicts the assumption 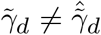. Therefore, the parameter 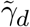 is uniquely identified. Furthermore, the equation 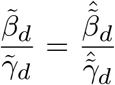 yields 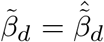. Similarly, 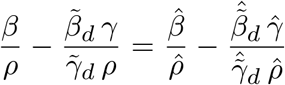 yields 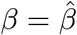.

## 4 Parameter Estimation and Practical Identifiability

### 4.1 Multiscale Model Fitting

In this study, we use a sequential fitting strategy [31, 34] to estimate parameters in the multiscale HIV model that combines within-host (microscale) and between-host (macroscale) dynamics. This approach leverages individual-level infection dynamics data to inform population-level epidemic parameters, ensuring biological consistency across scales. In the first stage of sequential fitting, the within-host model is fitted to viral load and total target cell counts from 80 HIV-infected patients (see Section 4.1.1), to estimate the within-host parameters (*λ, β*_*v*_, *δ*_1_, *δ*_2_, *p, c*). In the second stage, these within-host parameter estimates are incorporated into the between-host model, which is then fitted to population-level surveillance data to estimate the between-host parameters 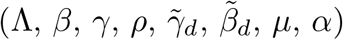 (see Section 4.1.2).

#### 4.1.1 Within-Host Model Fitting

We estimate the parameters of the within-host model (1), which describes the dynamics of target cells *T* (*τ*), infected cells *T*_*i*_(*τ*), and viral load *V* (*τ*), in an infected individual using nonlinear mixed-effect models. Nonlinear mixed-effect (NLME) modeling is a statistical approach used to analyze complex biological data, including pharmacokinetics and infectious disease dynamics [41]. The parameter set 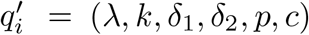 is estimated from 80 HIV-infected individuals enrolled in the AIDS clinical study [39]. Target cell counts and viral load measurements were collected from patients undergoing treatment with NRTI and PI. Because the data include repeated measures from multiple individuals, we employ a nonlinear mixed-effects (NLME) modeling framework to account for both fixed effects and random effects. Let *x*(*τ*) = (*T* (*τ*), *T*_*i*_(*τ*), *V* (*τ*)) represent the state variables of the within-host model (1). The observed quantities are defined as:

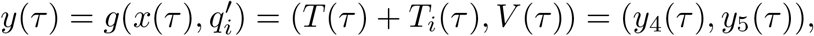

where 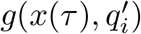 maps the state variables to the observed total CD^4+^ T-cell count and viral load. The relationship between the observations *y*_4_(*τ*), *y*_5_(*τ*) and the measured data is given by the following statistical model;

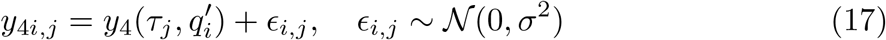

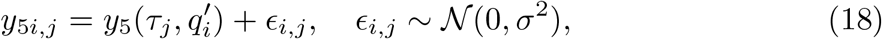

where 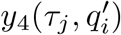 is the model prediction of total CD^4+^ T-cell counts (*T* (*τ*) + *T*_*i*_(*τ*)) at time *τ*_*j*_ for the *i*^*th*^ individual, and *y*_4*i,j*_ is the measured total CD^4+^ T-cell count in the clinical study. Similarly, 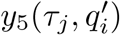 is the model prediction of the viral load *V* (*τ*) at time *τ*_*j*_ for the *i*^*th*^ individual, and *y*_5*i,j*_ is the measured viral load data. The within-host parameters for the individual *i* are modeled as

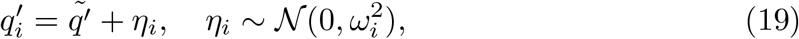

where 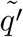 is the population mean (fixed effect) and *η*_*i*_ represents the individual-level random effect. Thus, each 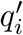 follows a normal distribution with mean 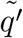 and standard deviation *ω*_*i*_. We use Monolix, a leading tool for NLME modeling [42], which is widely used in pharmacometrics and drug development. Monolix employs the Stochastic Approximation-Expectation Maximization (SAEM) algorithm [43] to estimate population parameters without requiring model approximation. It also supports model selection, diagnostics, and individual parameter estimation [**?**]. Pre-estimated initial conditions for the model are:

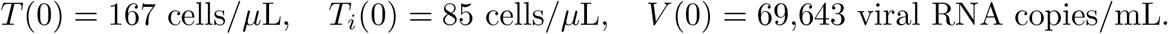

Figure 1 shows the distribution of within-host model parameter estimates from Monolix using nonlinear mixed-effects fitting. The estimated fixed effects 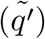 and their standard deviations (*ω*_*i*_) are presented in Table 1. The dynamics of uninfected and infected T cells, total T cells, and viral load for 80 patients are shown in Figure 2.

**Table 1.** Population parameter estimates and inter-individual variability from NLME fitting of the within-host model to data from 80 HIV-infected individuals.

| Parameter | Population Mean ($\tilde{q}'$) | Std Dev ($\omega_i$) | Unit |
| --- | --- | --- | --- |
| $\lambda$ | 33.84 | 0.40 | cells $\mu\text{L}^{-1}$ week<sup>-1</sup> |
| $\beta_v$ | $8.7 \times 10^{-3}$ | 0.89 | mL (RNA copies)<sup>-1</sup> week<sup>-1</sup> |
| $\delta_1$ | $3.6 \times 10^{-4}$ | 2.70 | week<sup>-1</sup> |
| $\delta_2$ | 0.085 | 0.56 | week<sup>-1</sup> |
| $p$ | 0.45 | 1.35 | $10^3$ RNA copies cell<sup>-1</sup> week<sup>-1</sup> |
| $c$ | 0.96 | 0.61 | week<sup>-1</sup> |

**Fig. 2.**
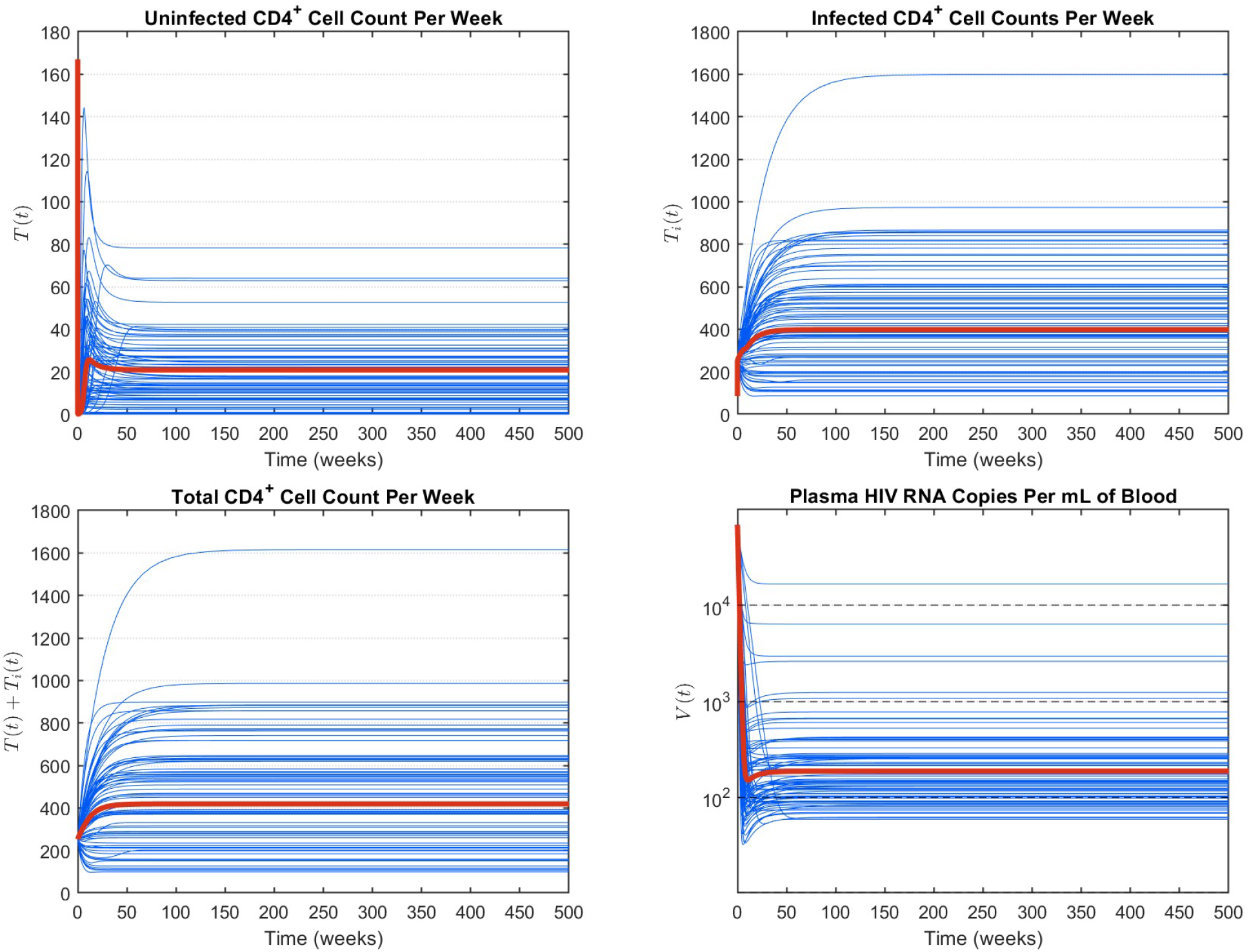
Dynamics of uninfected and infected T cells, total T cells, and viral load over time for 80 treated patients. Each blue line shows the trajectory for an individual patient using that patient’s SAEM-estimated parameters, capturing inter-patient heterogeneity in the target cell population and viral suppression under ART. The orange line denotes the population-level prediction based on the fixed effects.

The CD4^+^ T cell production rate was estimated as *λ* = 33.84 cells per *µ*L of blood per week. The estimated death rates were *δ*_1_ = 3.6 *×* 10^−4^ per week for uninfected CD4^+^ T cells, *δ*_2_ = 0.085 per week for infected cells. The clearance rate of the free virus is estimated at *c* = 0.96 per week. The infection rate of target cells by the virus, 16 *β*_*v*_ = 8.7 × 10^−3^ (mL·vRNA^−1^ · week^−1^), and the viral production rate per infected cell, *p* = 0.45 (10^3^ vRNA copies · cell^−1^ · week^−1^), are both expected to be reduced compared with untreated infection, since all patients in this cohort were receiving combination antiretroviral therapy (ART) including nucleoside reverse transcriptase inhibitors (NRTIs) and protease inhibitors (PIs). NRTIs block reverse transcription, while PIs prevent viral maturation, thereby jointly reducing *β*_*v*_ and *p*. The basic reproduction number, 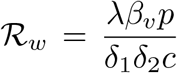, calculated for each individual in the HIV clinical study, was significantly larger than 1.

#### 4.1.2 Between-Host Model Fitting

The within-host model provides viral dynamics under treatment, with the estimated population means of within-host parameters (see Table 1) providing average viral load trajectories for HIV-infected individuals undergoing treatment. These trajectories are then used as inputs to the between-host epidemic model, establishing a link between individual-level viral kinetics with transmission and the progression to AIDS at the population scale. In the second stage of the sequential fitting procedure, we validate the between-host model using surveillance data. Specifically, we estimate the values of the between-host parameters 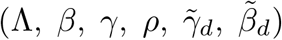 by fitting the epidemic model to three data sets: annual HIV incidences, AIDS classifications, and HIV diagnoses obtained from the CDC AtlasPlus database [36], which correspond *y*_1_(*t*), *y*_2_(*t*), and *y*_3_(*t*), respectively.

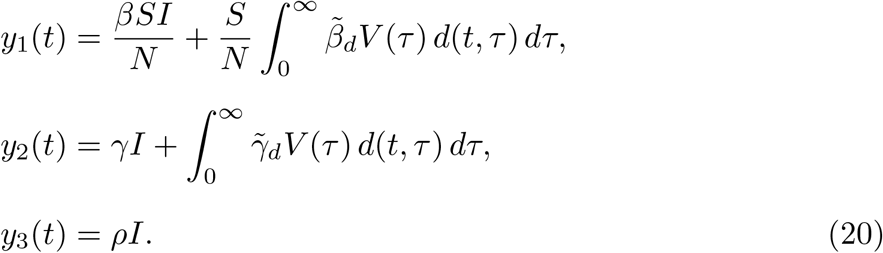

where the viral load *V* (*τ*) is determined from the population-level prediction of the within-host model (see Table 1). The initial conditions are pre-estimated and then set to the values, *S*(0) = 820, *I*(0) = 42, *A*(0) = 0.337, and *d*(0, *τ*) = 0 for all *τ*. Since the multiscale model is structurally identifiable, when the AIDS-related death rate is known, we set *α* = 0.002 year^−1^. Furthermore, the natural death rate is fixed at *µ* = 0.02 year^−1^. The system is numerically integrated using a finite difference scheme. Parameters are estimated by minimizing the following weighted least-squares objective function using MATLAB’s *fmincon* algorithm,

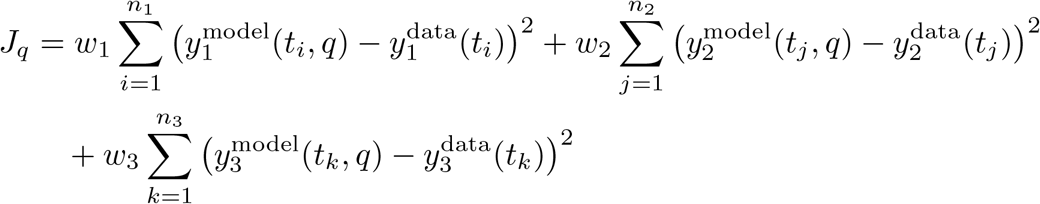

with constraint

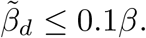

This constraint ensures that treatment reduces the transmission rate. The weights in the least-squares objective function are *w*_1_ = 1 for HIV incidence, *w*_2_ = 10 for AIDS classifications, and *w*_3_ = 20 for HIV diagnoses. The parameter bounds and the estimated values are reported in Table 2. The estimate for 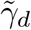 was extremely small (2.44 *×* 10^−16^), suggesting negligible AIDS progression for treated individuals. Based on this result, we refit the model by fixing 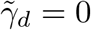 and adjusting the weights to *w*_1_ = 2, *w*_2_ = 11, and *w*_3_ = 20. Table 2 summarizes the final parameter estimates for both model assumptions, with and without fixing 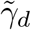. Figure 3 presents the results of the model fit against the CDC-reported data, including the estimated annual incidence of HIV, annual diagnoses, and the classification of AIDS cases in the United States.

**Table 2.**
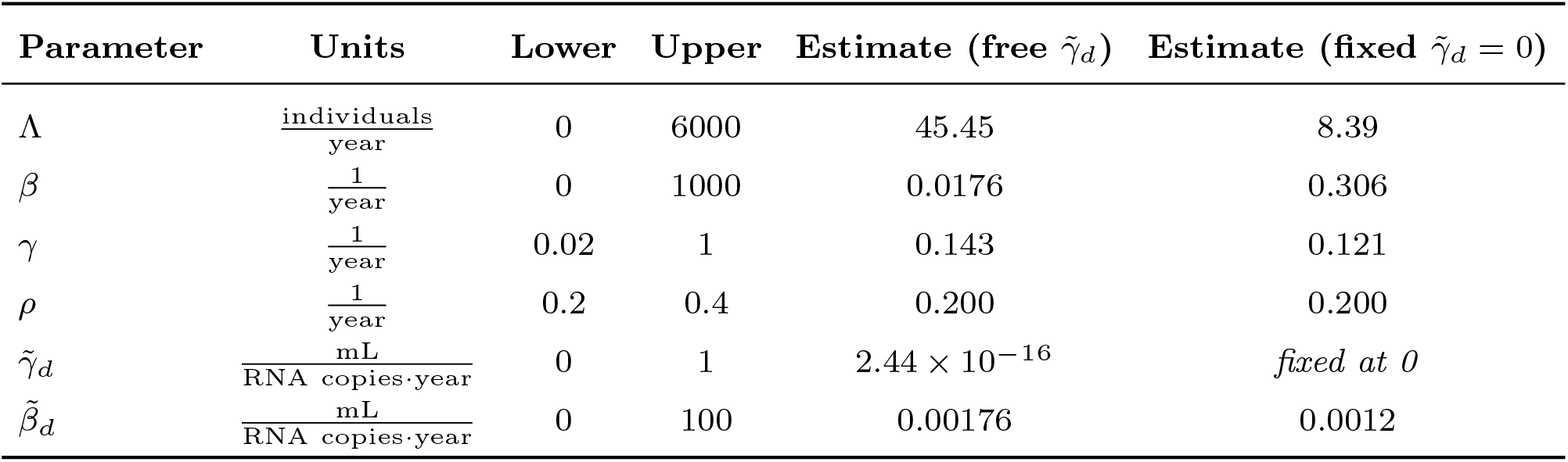
Estimated parameter bounds and values for the between-host HIV model, comparing the cases with and without fixing 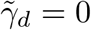.

| Parameter | Units | Lower | Upper | Estimate (free $\tilde{\gamma}_d$) | Estimate (fixed $\tilde{\gamma}_d = 0$) |
| --- | --- | --- | --- | --- | --- |
| $\Lambda$ | $\frac{\text{individuals}}{\text{year}}$ | 0 | 6000 | 45.45 | 8.39 |
| $\beta$ | $\frac{1}{\text{year}}$ | 0 | 1000 | 0.0176 | 0.306 |
| $\gamma$ | $\frac{1}{\text{year}}$ | 0.02 | 1 | 0.143 | 0.121 |
| $\rho$ | $\frac{1}{\text{year}}$ | 0.2 | 0.4 | 0.200 | 0.200 |
| $\tilde{\gamma}_d$ | $\frac{\text{mL}}{\text{RNA copies} \cdot \text{year}}$ | 0 | 1 | $2.44 \times 10^{-16}$ | *fixed at 0* |
| $\tilde{\beta}_d$ | $\frac{\text{mL}}{\text{RNA copies} \cdot \text{year}}$ | 0 | 100 | 0.00176 | 0.0012 |

**Fig. 3.**
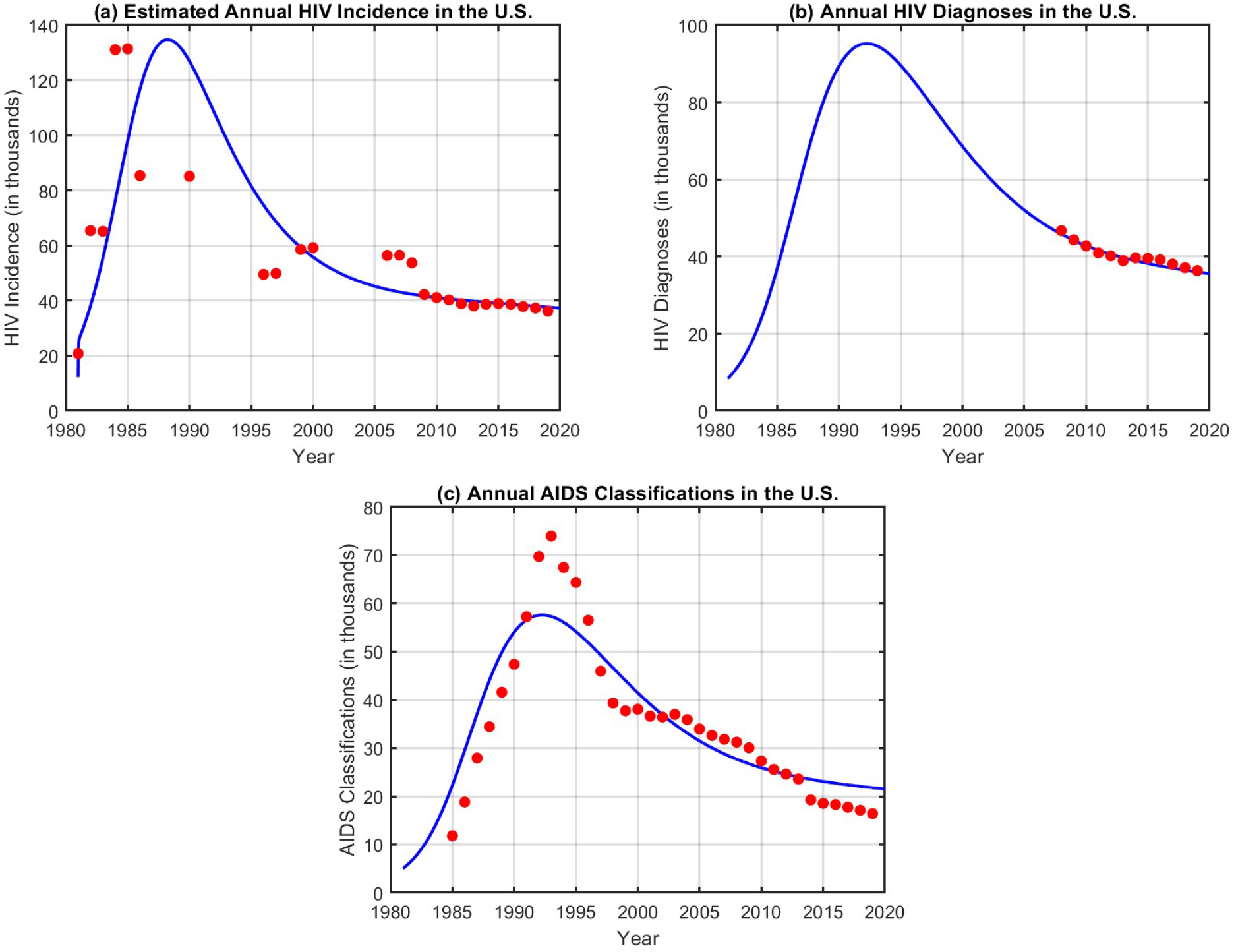
Model fitting results against CDC data in the United States. **(a)** Estimated annual HIV incidence from 1981 to 2020. Red circles correspond to CDC-reported data in Table B3. **(b)** Annual HIV diagnoses reported from 2008 to 2020 (Table B5). **(c)** AIDS case classifications from 1985 to 2020 (Table B4). In all panels, blue curves represent model predictions.

### 4.2 Practical Identifiability Analysis of the Multiscale HIV Model

Practical identifiability addresses challenges associated with real-world data, which are often noisy, incomplete, or limited. Even when a model is structurally identifiable, parameter estimates obtained through numerical optimization may still be unreliable due to non-uniqueness under noisy conditions. Therefore, both structural and practical identifiability must be considered when fitting models to data. In this study, we use the Monte Carlo simulation (MCS) approach to assess the practical identifiability of the multiscale HIV model [27]. We provide the details of the MCS in Appendix E. To interpret the MCS results, we evaluate the practical identifiability of each parameter based on its calculated *ARE*, as defined below [27].

**Definition 3** The practical identifiability of a parameter *q* is determined by comparing its average relative estimation error *ARE*(*q*) to the measurement error *σ*:

1. If 0 ≤ *ARE*(*q*) ≤ *σ*, then the parameter *q* is strongly practically identifiable.
2. If *σ < ARE*(*q*) ≤ 10*σ*, then the parameter *q* is weakly practically identifiable.
3. If *ARE*(*q*) *>* 10*σ*, then the parameter *q* is not practically identifiable.

To evaluate the practical identifiability of parameters in the between-host model, we conduct two separate Monte Carlo simulations based on two distinct parameter sets. These sets are obtained by fitting the model to data under two scenarios: (1) estimating all parameters, including 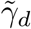, and (2) fixing 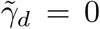 due to its negligible influence on the model fit. The two fittings result in different parameter values. Using these fitted sets as references, we simulate synthetic datasets under varying levels of noise (*σ* = 1%, 5%, 10%, 20%), re-estimate the parameters, and compute the average relative estimation error (ARE) for each parameter in both scenarios.

Table 3 summarizes the average relative estimation errors for each parameter under different noise levels, comparing the estimates with and without fixing 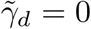. When 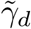 is freely estimated, it is not practically identifiable, with ARE values exceeding 10^11^ even at the lowest noise level. The parameter *β* becomes non-identifiable across all noise levels, with ARE values ranging from 18.7% to more than 400%. The parameter 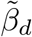 is weakly identifiable at all noise levels except at *σ* = 10%, where it is strongly identifiable. In contrast, the parameters Λ, *γ* and *ρ* are strongly practically identifiable at all noise levels tested. When 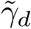 is fixed to zero, the identifiability of the remaining parameters improves substantially. The parameter *β* is strongly practically identifiable across all noise levels, with ARE values consistently below the corresponding *σ* threshold. Similarly, 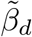 maintains strong identifiability under all conditions, even at *σ* = 20%, where its ARE remains under 10%. The parameters *γ* and *ρ* are strongly identifiable at all noise levels. The parameter Λ does not achieve strong identifiability at any noise level. Its ARE slightly exceeds *σ* in 1% noise and remains between *σ* and 10*σ* at higher noise levels, indicating weak practical identifiability throughout.

**Table 3.** Monte Carlo simulation results: Absolute Relative Estimation Error (ARE) for each parameter across varying noise levels, comparing estimates with and without fixing 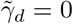.

| Noise Level | ARE (Free $\tilde{\gamma}_d$) | | | | | | ARE (Fixed $\tilde{\gamma}_d = 0$) | | | | |
| --- | --- | --- | --- | --- | --- | --- | --- | --- | --- | --- | --- |
| | $\Lambda$ | $\beta$ | $\gamma$ | $\rho$ | $\tilde{\gamma}_d$ | $\tilde{\beta}_d$ | $\Lambda$ | $\beta$ | $\gamma$ | $\rho$ | $\tilde{\beta}_d$ |
| $\sigma = 1\%$ | 0.20 | 18.70 | 0.65 | 0.71 | $1.9 \times 10^{11}$ | 1.15 | 1.23 | 0.44 | 0.59 | 0.49 | 0.57 |
| $\sigma = 5\%$ | 1.09 | 88.63 | 3.47 | 3.82 | $1.26 \times 10^{12}$ | 5.43 | 6.24 | 2.22 | 2.81 | 2.29 | 2.74 |
| $\sigma = 10\%$ | 2.33 | 156.59 | 6.98 | 8.00 | $2.69 \times 10^{12}$ | 9.84 | 12.87 | 4.73 | 6.19 | 5.37 | 5.47 |
| $\sigma = 20\%$ | 6.45 | 406.74 | 14.53 | 15.75 | $4.95 \times 10^{12}$ | 20.09 | 27.22 | 9.76 | 13.54 | 11.81 | 9.98 |

## 5 Can We End the HIV Epidemic in the USA?

In 2019, the United States launched the Ending the HIV Epidemic (EHE) initiative, an ambitious national program aimed at reducing new HIV infections by 90% by 2030. The initiative relies on four primary intervention strategies: diagnose, treat, prevent, and respond. Early diagnosis reduces the time individuals remain unaware of their infection and capable of transmitting the virus. Immediate initiation of antiretroviral therapy with sustained viral suppression will effectively remove treated individuals from the HIV transmission process. In parallel, the widespread use of pre-exposure prophylaxis (PrEP) can reduce the risk of infection among susceptible populations. Although PrEP is a cornerstone of EHE prevention efforts and has been shown to reduce the risk of HIV acquisition by up to 99% among high-risk individuals, our multiscale model does not explicitly incorporate PrEP uptake. We analyze the impact of PrEP within the EHE initiative in a separate study [44]. EHE aims to reduce new infections to 9,300 by 2025 and to 3,000 by 2030. The initiative sets goals of 9,600 diagnoses by 2025 and 3,000 by 2030, ensuring that at least 95% of people living with HIV are aware of their status. The HIV epidemic in the United States began in 1981, with incidence rising sharply through the early 1980s, peaking in the mid-1980s, and then gradually declining through the 1990s and 2000s. We fitted the multiscale model to HIV surveillance data from 1981 to 2019, covering both the early years of the epidemic and the period leading up to the Ending the HIV Epidemic (EHE) initiative. Figure 4 shows model projections through 2030 for the EHE pillars: HIV incidence, diagnoses, and the proportion of individuals aware of their HIV status. In our multiscale model, the knowledge of HIV status is represented as 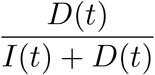 where *I* represents undiagnosed infections and *D* diagnosed cases. This outcome was not directly fitted to the data but simulated to assess progress towards the (EHE) target of 95% awareness of HIV status by 2025. In addition to these epidemiological outcomes, the baseline parameter set yields a basic reproduction number of ℛ_0_ = 4.037. The epidemic threshold is given by 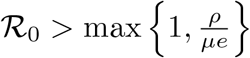, which accounts for both natural removal and the rate of diagnosis. With *ρ* = 0.2 and *µ* = 0.02, we obtain *ρ/*(*µe*) ≈ 3.68. Since ℛ_0_ ≈ 4.037 exceeds this threshold, the model predicts that, without EHE interventions, HIV transmission would have been sustained in the population, as seen in Figure 4. Next, we consider one of the key indicators of the EHE initiative: sustained viral suppression. In this scenario, we assume that diagnosed individuals achieve complete viral suppression beginning in 2019 with the start of EHE initiative, and set 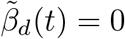 for *t >* 2019. Figure 5 presents model projections under this assumption. The projections show a rapid decline in HIV incidence, highlighting the critical role of sustained viral suppression in reducing new infections. The number of diagnoses also declines, although not as steeply as incidence, and falls to levels broadly consistent with the EHE targets for 2025 and 2030. If viral load could have been reduced below the detectable threshold for all treated individuals beginning in 2019, then the model projects that incidence and awareness goals would have been achieved by 2025 and sustained through 2030, while diagnosis levels would approximate national targets. This scenario also produces a sharp reduction in the basic reproduction number (Figure 5d), which drops below the epidemic threshold after 2019 (ℛ_0_ ≈ 0.898), confirming that HIV transmission cannot be sustained in the absence of infectiousness from diagnosed individuals.

**Fig. 4.**
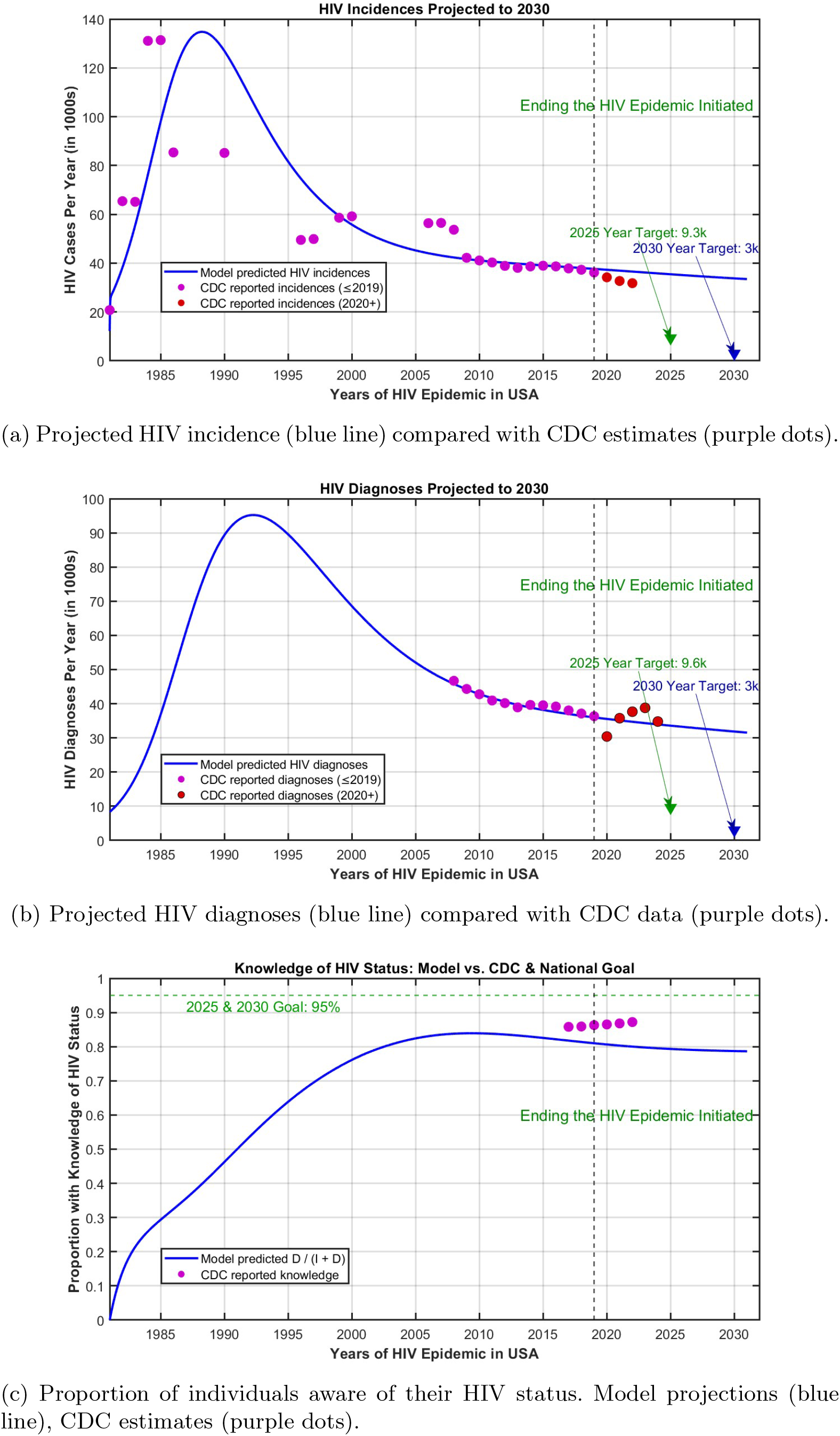
Blue lines are model projections, purple dots are CDC data through 2019 used for fitting, and red dots are CDC data from 2020 onward used for validation. The vertical dashed line marks the 2019 launch of the Ending the HIV Epidemic initiative, and green arrows indicate the national targets for 2025 and 2030. Panel (a) shows projected HIV incidence, panel (b) shows projected HIV diagnoses. Panel (c) shows knowledge of HIV status and model output is compared with CDC data and the 95% national goal.

**Fig. 5.**
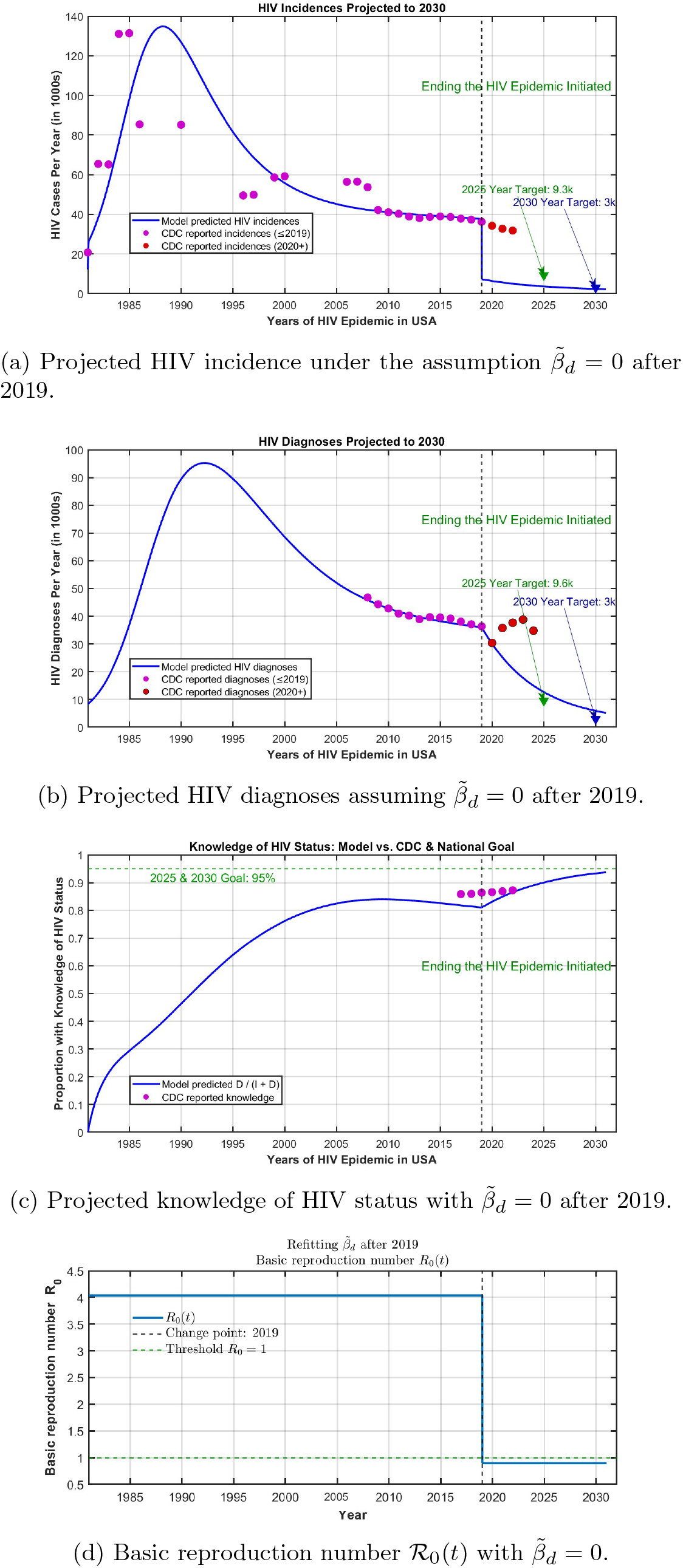
Model projections under a hypothetical scenario where transmission from diagnosed individuals is set to zero 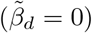 after 2019. Blue lines are model projections, purple dots are CDC data through 2019 used for fitting, and red dots are CDC data from 2020 onward used for validation. The vertical dashed line marks the 2019 launch of the Ending the HIV Epidemic initiative, and green arrows indicate the national targets for 2025 and 2030. Panel (a) shows HIV incidence, panel (b) shows HIV diagnoses, and panel (c) shows knowledge of HIV status, model output is compared with CDC data and the 95% national goal. Panel (d) shows the basic reproduction number ℛ_0_(*t*), which drops below the epidemic threshold once diagnosed transmission is removed.

It is not realistic to assume immediate success at viral suppression at the start of the initiative. Therefore, we next consider a scenario in which transmission from diagnosed individuals decreases gradually over time. Figure 6 presents model projections when 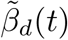 is refitted as a time-dependent parameter for the period 2019-2030 to align with EHE goals. Prior to 2019, 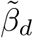 remains constant at its fitted value of 0.0012, while after 2019 it decreases linearly to meet the national targets. The fitted time-dependent transmission parameter 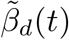 is given by,

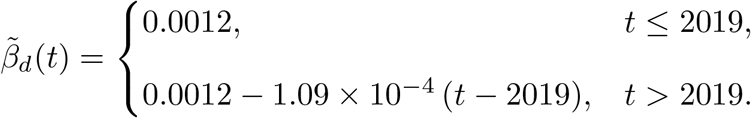

**Fig. 6.**
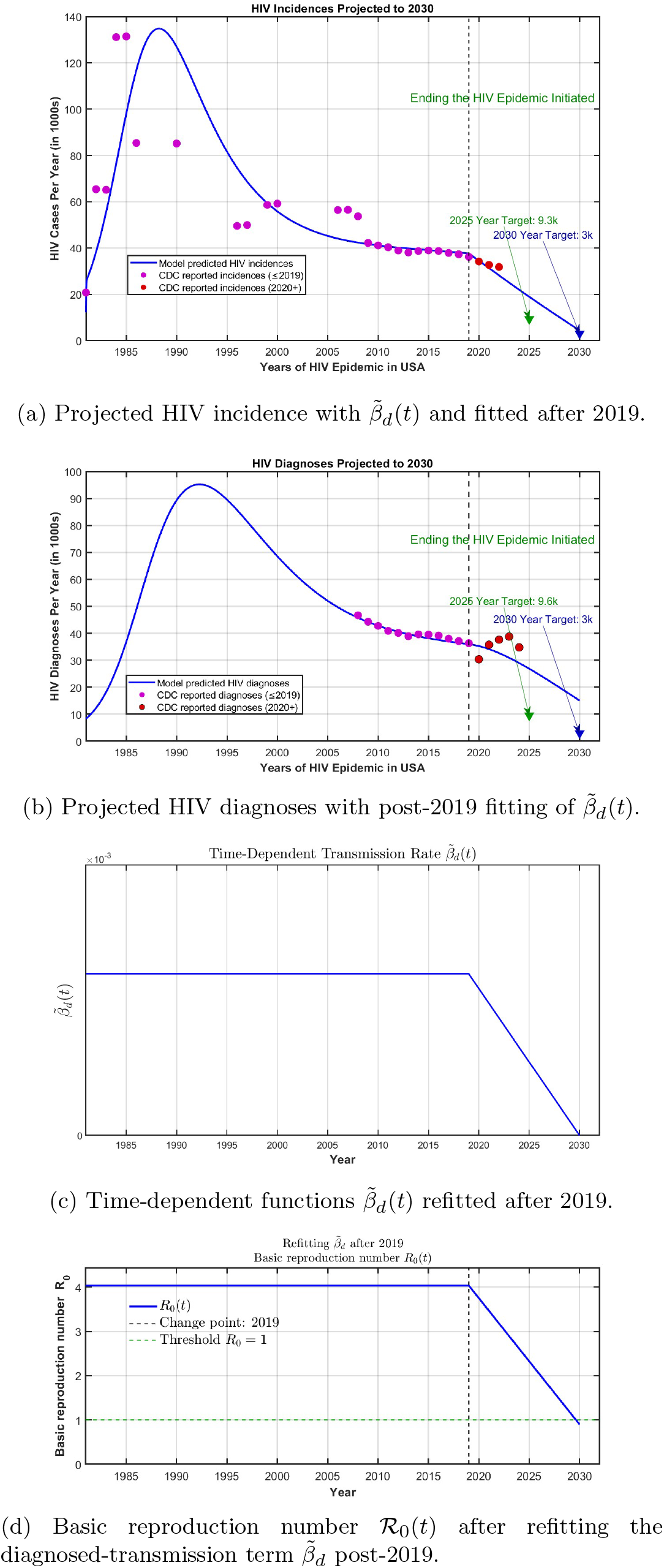
Model projections with a time-varying transmission rate from diagnosed individuals, 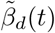, refitted after 2019. Blue lines are model projections, purple dots are CDC data through 2019 used for fitting, and red dots are CDC data from 2020 onward used for validation. The vertical dashed line marks the 2019 launch of the Ending the HIV Epidemic initiative, and green arrows indicate the national targets for 2025 and 2030. Panels (a) and (b) show projected incidence and diagnoses, panel (c) shows the refitted function 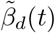, and panel (d) shows the corresponding basic reproduction number ℛ_0_(*t*).

Under this scenario, HIV incidence declines after 2019 and meets the 2030 EHE incidence goal, although the 2025 target is not achieved. The number of diagnoses also decreases, but more gradually, and remains above the EHE diagnosis targets for both 2025 and 2030. The reproduction number follows a parallel downward trend: ℛ_0_ = 4.037 in 2018, drops to ℛ_0_ ≈ 2.32 by 2025, and falls below the epidemic threshold with ℛ_0_ ≈ 0.898 in 2030. These results suggest that reductions in infectiousness among diagnosed individuals alone are sufficient to bring incidence under control by 2030, but additional improvements in diagnosis rates are needed to meet the diagnosis-related EHE goals.

Furthermore, we explored the impact of increasing the diagnosis rate on the EHE goals and considered a combined intervention scenario in which transmission from treated individuals was reduced while the diagnosis rate was increased. Our analysis indicates that increasing the diagnosis rate *ρ*(*t*) alone is not sufficient to meet the EHE targets (results not shown), suggesting that improved efforts at diagnosis alone cannot drive the epidemic decline at the required pace. In the combined intervention scenario, we observe results similar to those obtained when only transmission from treated individuals is reduced (see Figure F2). Specifically, HIV incidence continued to decline and reached the 2030 EHE incidence target, although the 2025 target was not achieved. However, the number of diagnoses declined more slowly and did not meet either the 2025 or the 2030 diagnosis targets, indicating that reductions in transmission primarily affect incidence, while diagnosis-related targets are more difficult to achieve.

### Impact of within-host patient variability on the epidemiological surveillance data

Parameter estimation for the HIV multiscale model was carried out by fixing within-host parameters at their population mean values (see Table 1). It is clearly evident from Figure 2, there exists substantial inter-individual variability among the 80 patients in both CD4^+^ T cell counts and HIV viral load. The next numerical experiment investigates how this variability is transmitted to population-level surveillance dynamics. Each patient is associated with a distinct within-host viral load trajectory, which is then projected to the epidemic scale through the coupled multiscale model. This approach transfers individual variability into population-level predictions. Consequently, the resulting ensemble of epidemic trajectories reflects how heterogeneity in viral dynamics across patients propagates into variability in key EHE indicators, including HIV incidence and diagnoses (see Figure 7). We see that variation in viral load among patients is directly transmitted to the population model, resulting in a broad spread of epidemic trajectories. The ensemble highlights how strongly individual heterogeneity can amplify uncertainty in epidemic-scale predictions.

**Fig. 7.**
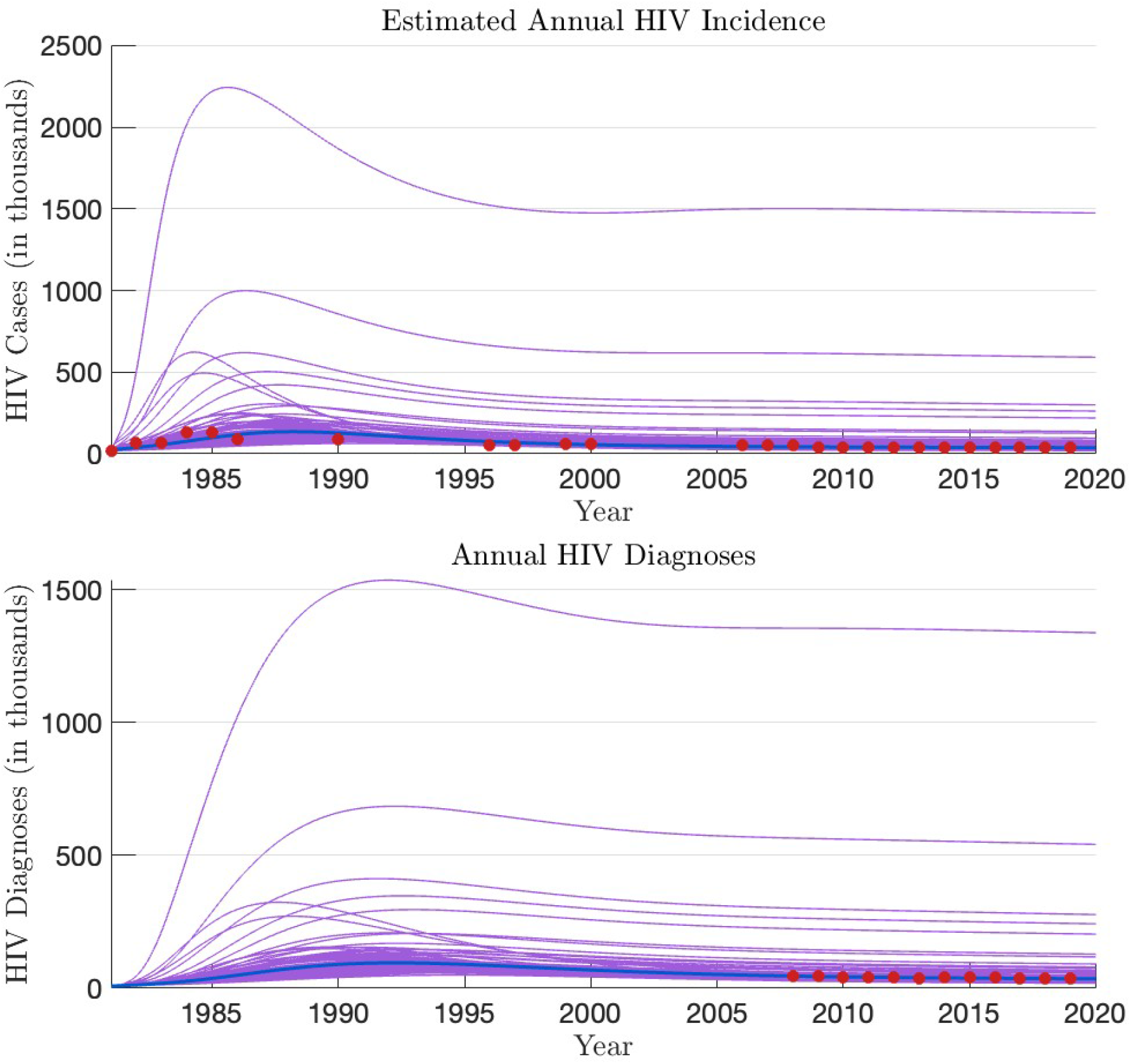
Thin purple curves show simulations based on 80 patient-specific within-host parameter sets, whereas the thick blue curve corresponds to the simulation obtained using population mean values. Red markers indicate national surveillance data (in thousands) and the blue curve represents the model fit to these data.

## 6 Virtual Epidemics

In this section, we introduce the concept of virtual epidemics, inspired by the idea of virtual patients, that has been widely used in pharmacokinetics to evaluate treatment strategies [45–48]. A virtual patient is a parameterization of a mathematical model that captures inter-individual (patient) variability while remaining consistent with physiological and clinical constraints [45, 47, 48]. Collections of such parameterizations define virtual patient populations, which help researchers explore disease progression differences and responses to therapies. We extend this framework to infectious disease dynamics and define virtual epidemics as parameterizations of the multiscale model (1). In other words, both the within-host and between-host models are parameterized to capture individual- and population-level variability while remaining epidemiologically feasible. By generating virtual epidemics, we can examine transmission variability, evaluate interventions, and test prediction robustness under parameter uncertainty.

The construction of virtual patient populations involves the following key steps. The process starts with the formulation of the mathematical model and then the model parameters are estimated using data from existing biological studies, laboratory experiments, or clinical trials. Next, the sensitivity of the model predictions to perturbations in parameter values is determined to identify influential parameters. Finally, virtual patients are generated using one of the two common approaches: (a) **Sampling-based approach:** parameter values are drawn at random from predefined distributions, and a candidate parameter set is accepted if the resulting model predictions are within the acceptable deviations from the known outcomes and rejected otherwise [47, 48], or (b) **Data fitting approach:** virtual patients are obtained directly from successful model calibrations; parameters that fit the model to data using an optimization algorithm are automatically incorporated into the virtual population [45, 48]. We take the data-fitting approach to generate virtual epidemics.

Epidemiological surveillance data, such as reported HIV incidence rates, offer valuable information for public health; however, they have substantial limitations when used in mathematical models aimed at forecasting future disease trajectories. The main issues with epidemic data involve underreporting, delays in reporting, and changing diagnostic criteria or testing policies, which introduce significant uncertainty and compromise the reliability of predictions. Surveillance data only give an imperfect picture of the real epidemic. They provide a noisy approximation of the true epidemic. Motivated by these limitations, we propose the idea of virtual epidemics. Surveillance data provide a single instance of an ongoing epidemic, and with virtual epidemics, we generate several instances to better understand the true epidemic. The construction of virtual epidemics proceeds with the following steps. First, we use the available epidemic data to estimate model parameters. At this stage, we ensure these parameters are both structurally and practically identifiable. Only the parameters that satisfy these criteria are used to generate virtual epidemics. We proceed with creating virtual epidemics that preserve epidemiological realism, such as having epidemic trajectories that align closely with observed dynamics. To achieve this, we first compute the standard deviations of the surveillance data, then generate datasets that lie within three standard deviations of the empirical mean. These data sets are then fitted to the model to produce new epidemic trajectories. These trajectories, referred to as virtual epidemics, are possible scenarios of epidemic dynamics that are consistent with observed surveillance data.

Figure 8 shows the probability distributions of the parameters Λ, *β, γ, ρ*, and 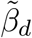 obtained from 5000 virtual epidemic trajectories, trimmed to retain the central 99% (0.5–99.5 percentiles). The panels illustrate parameter uncertainty and variability across the virtual epidemic trajectories: Λ, *β* and 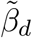 exhibit normal distributions; *γ* is right-skewed with a longer upper tail; *ρ* is strongly concentrated near its lower bound.

**Fig. 8.**
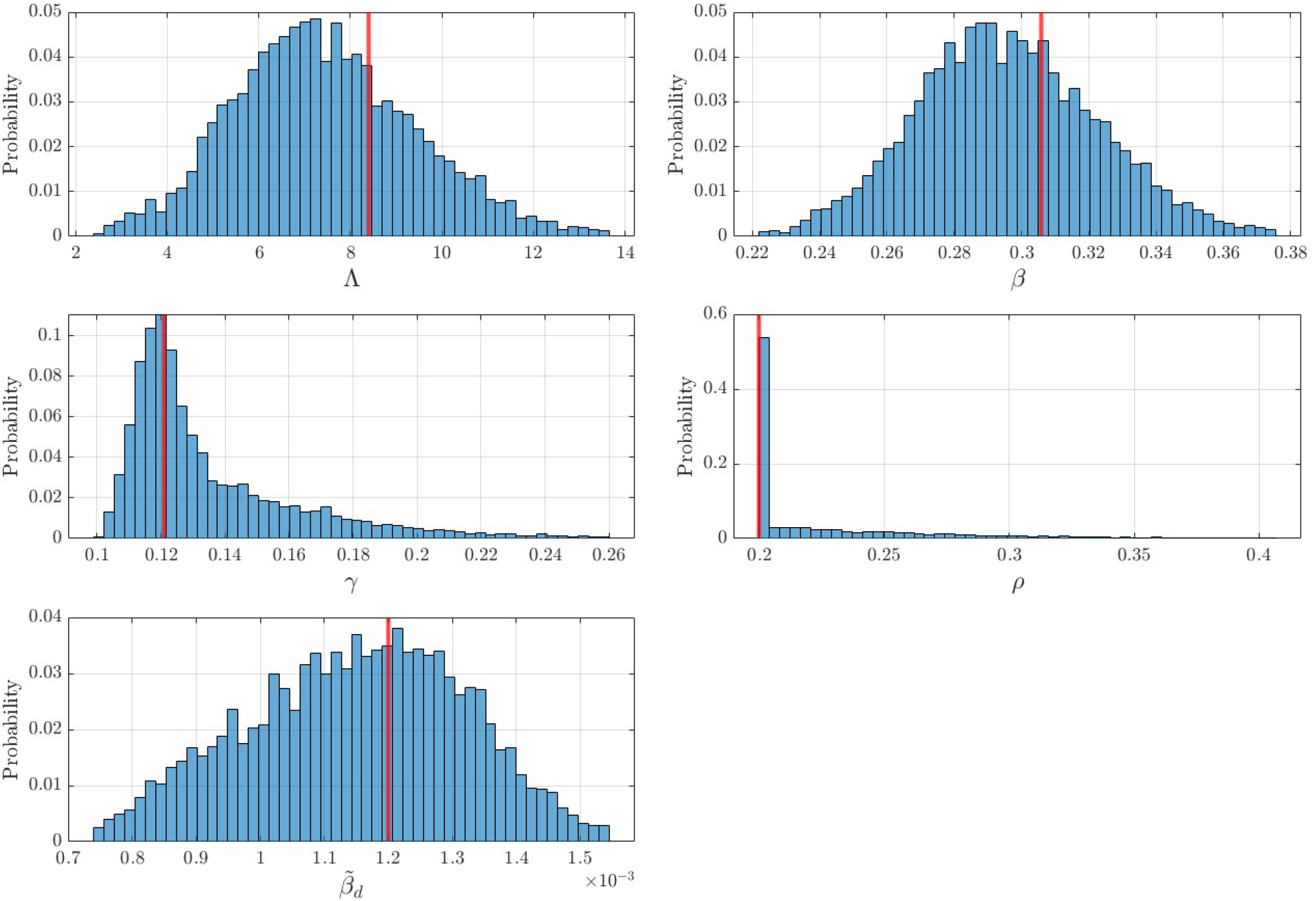
Histograms of trimmed bootstrap distributions for the parameters Λ, *β, γ, ρ*, and 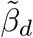, based on 5,000 bootstrap samples. For each parameter, the central 99% of values (0.5th and 99.5th percentiles) are retained to exclude extreme outliers. Histograms are normalized to show probability distributions. The red vertical line indicates the corresponding fitted parameter value from the fixed 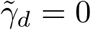 model reported in Table 2.

Figure 9 presents the results for the estimated annual HIV incidence, HIV diagnoses, and AIDS classifications in the United States. The curves represent the central 99% of virtual epidemics (0.5–99.5 percentiles). In each panel, thin teal lines correspond to individual epidemic trajectories, magenta points indicate data generated within the acceptable region which is within three standard deviations of the empirical mean at the CDC reporting years, the solid navy line denotes the best-fit model, and red circles show CDC-reported data for direct comparison.

**Fig. 9.**
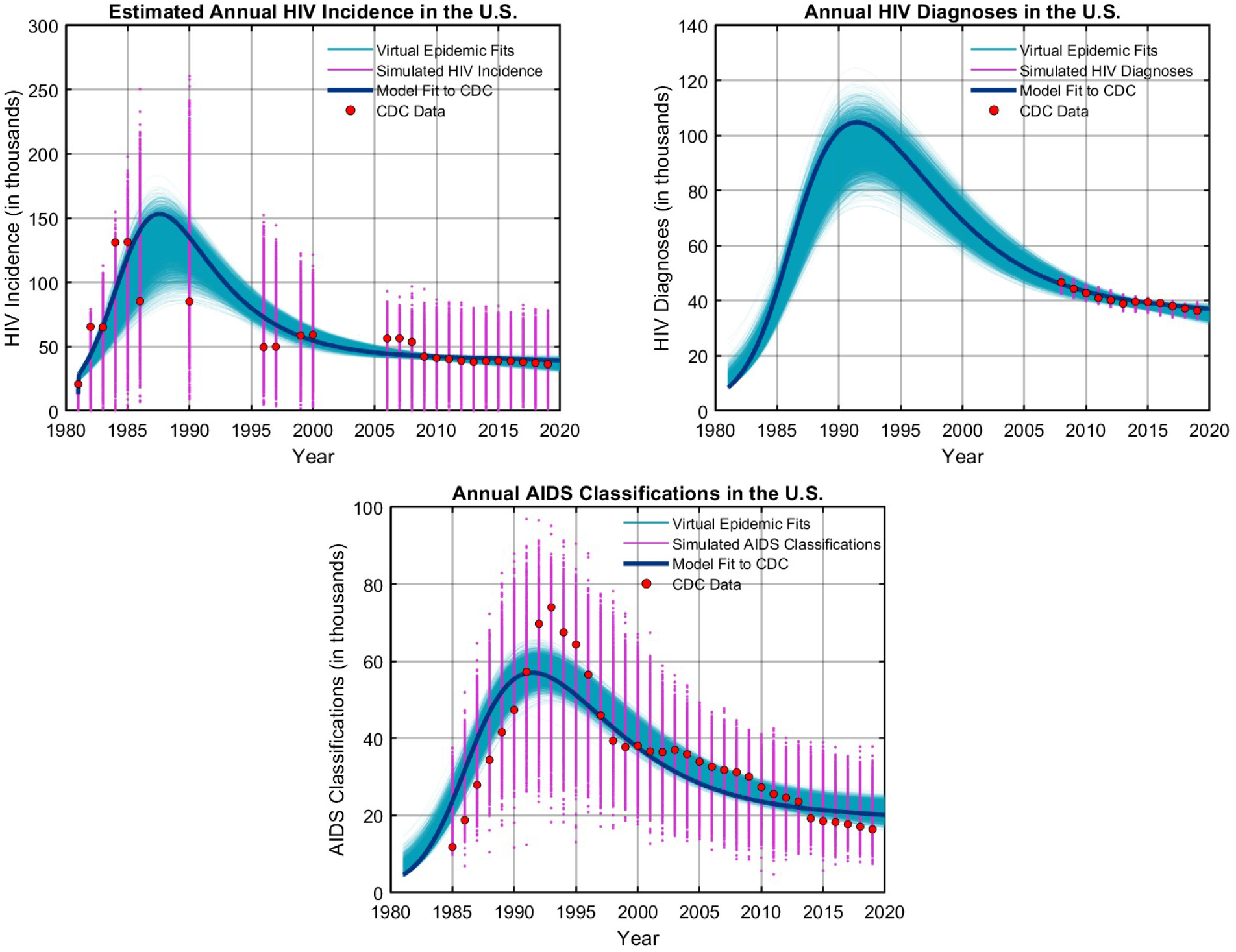
Virtual epidemic fits and model results for U.S. HIV data through 2020. Thin teal curves show an ensemble of virtual epidemic fits (*n* = 5000); magenta points mark ensemble predictions at CDC reporting years; the solid navy line shows the best-fit model; and red circles denote CDC-reported data.

**Fig. 10.**
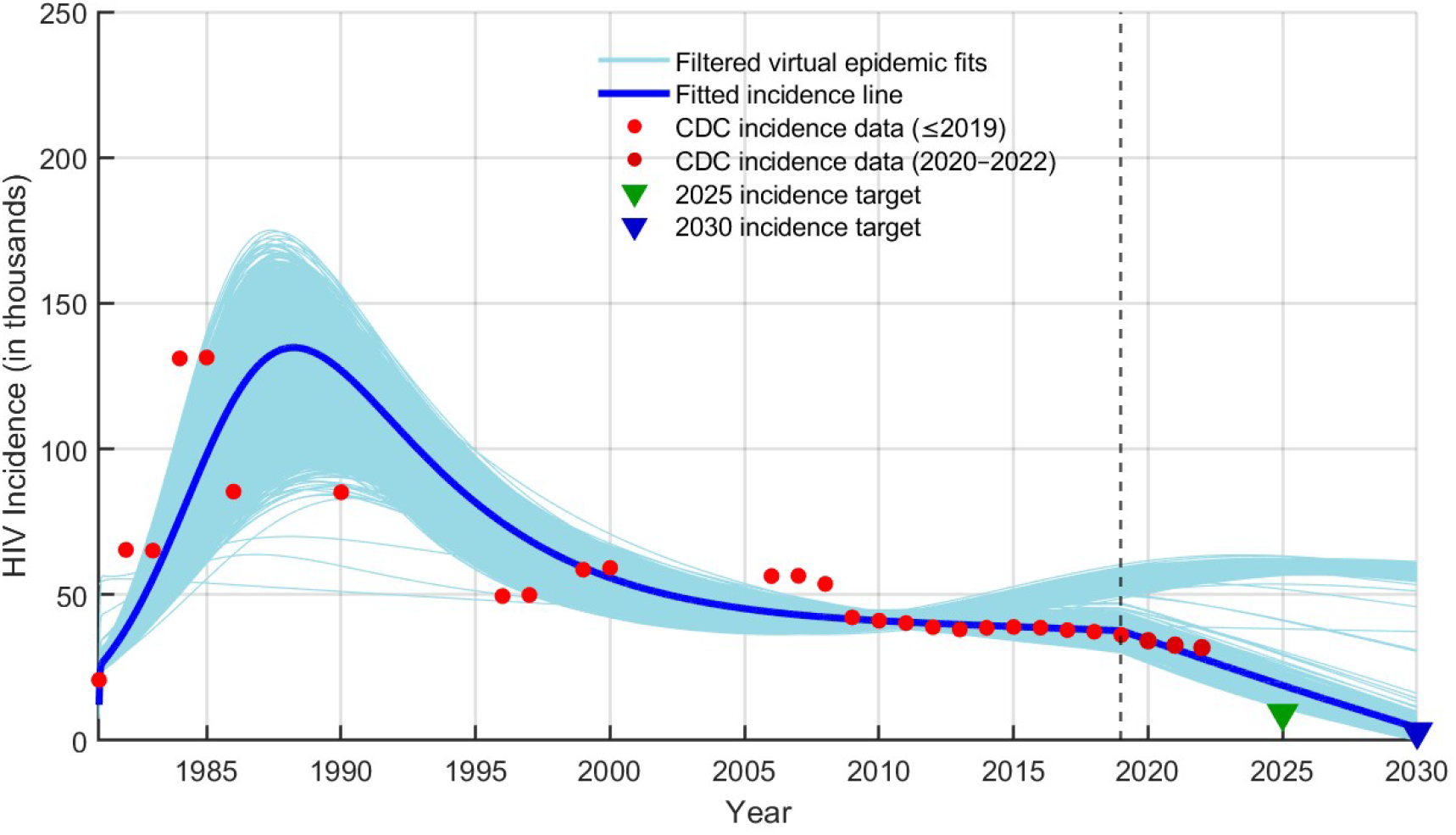
Filtered virtual epidemic projections of HIV incidence in the United States generated from 5,000 bootstrap parameter sets under a post-2019 decline in the transmission parameter 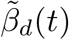. Virtual epidemic trajectories with unrealistically large incidence spikes (with maximum incidence exceeding 250,000 cases) were excluded for visualization purposes. The remaining trajectories are shown in light blue, while the solid blue curve represents the fitted incidence trajectory. Red markers denote CDC-reported HIV incidence data prior to 2019, and dark red markers indicate CDC-reported incidence data from 2020–2022. The green and blue markers denote the Ending the HIV Epidemic (EHE) incidence targets for 2025 and 2030, respectively. The dashed vertical line at 2019 marks the initiation of the EHE strategy and the onset of the linear decline in 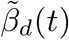 toward near-zero levels by 2030.

### Virtual Epidemic Predictions on Ending the HIV Epidemic

To quantify the probability that the Ending the HIV Epidemic (EHE) initiative can achieve its national HIV incidence targets under parameter uncertainty, we constructed a bootstrap ensemble of multiscale model parameter sets, each representing a virtual HIV epidemic. For each virtual epidemic, we fixed the core transmission and progression parameters (Λ, *β, γ, ρ*) and re-estimated only the post-2019 value of the treatment-related transmission modifier 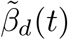, subject to epidemiologically plausible bounds, to favor the national goals of 9,300 new infections in 2025 and 3,000 in 2030. Each optimized virtual epidemic was projected forward to 2030, and we recorded whether the simulated incidence in 2025 and 2030 fell within *±*2,000 cases of the respective targets. Out of 5,000 virtual epidemics, only 0.04% reached the 2025 incidence target, whereas 67.08% reached the 2030 target. Under feasible post-2019 reductions in 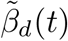, none of the virtual epidemics met both incidence targets simultaneously. These results suggest that, under parameter uncertainty, the long-term 2030 EHE incidence goal is substantially more achievable than the aggressive short-term reductions aimed for 2025. Although the 2025 target was rarely achieved in our simulations, the results indicate that sustained prevention and treatment efforts may still support progress toward the 2030 goal.

## 7 Conclusion

In this study, we have developed and analyzed a multiscale HIV model that directly couples within-host viral dynamics to population-level transmission and progression. By explicitly linking viral load to both infectiousness and disease progression and incorporating treatment age structure, the model captures key biological and clinical features of HIV epidemiology in the United States.

From a mathematical standpoint, we derived explicit threshold conditions governing the system’s long-term dynamics. The basic reproduction number ℛ_0_ was shown to be the primary determinant of persistence: when ℛ_0_ *<* 1, HIV is expected to die out, whereas ℛ_0_ *>* 1 ensures the existence of a positive endemic equilibrium. Furthermore, through a rigorous stability analysis, we established that the endemic equilibrium is locally asymptotically stable provided 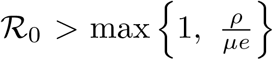. This condition highlights the interplay between biological processes (viral replication, diagnosis, and disease-induced mortality) and epidemiological thresholds, and provides a mathematically precise link between individual-level infection dynamics and the persistence of HIV at the population level.

A central methodological advance of this work is the analytical structural identifiability analysis, conducted directly on the coupled PDE-ODE system using differential algebra techniques. In the absence of established software for PDE-based models, we manually derived multiple input–output equations and demonstrated that all model parameters are structurally identifiable under realistic initial conditions and with the AIDS-induced death rate fixed. This establishes that parameter estimation from ideal data is theoretically well-posed.

Building on this, we implemented a sequential fitting approach: first estimating within-host parameters using patient-level viral load and CD4^+^ count data through a nonlinear mixed-effects (NLME) framework, then fitting the population-level model to CDC surveillance data on HIV incidence, diagnoses, and AIDS classifications. This approach was adopted because the within-host and population-level datasets have fundamentally different structures. The within-host model was fitted to data from 80 HIV-infected individuals using nonlinear mixed-effects modeling, which accounts for inter-individual variability in viral load trajectories, CD4^+^ counts, and observation times, whereas the population-level model was fitted to surveillance data using a least-squares framework. Epidemic trajectories were simulated using a finite difference method.

Our results demonstrate that addressing practical identifiability is essential when interpreting parameter estimates in multiscale epidemic models. While the full model is structurally identifiable, practical identifiability is sensitive to the inclusion of weakly informed parameters. In particular, allowing 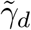(the treatment-dependent AIDS progression rate) to be estimated freely leads to severe non-identifiability not only for 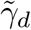 itself but also for key parameters such as *β* and 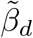. By fixing 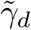 = 0, the identifiability of the remaining parameters improves substantially: 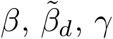, and *ρ* all become strongly practically identifiable across a range of noise levels. Only Λ, which enters the model indirectly through population recruitment, remains weakly identifiable.

Under baseline assumptions, the model reproduced surveillance trends but projected that the 2030 EHE goals for incidence and diagnoses would not be met. Scenario analyses revealed the central importance of transmission from diagnosed individuals. In the idealized case of complete viral suppression 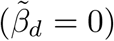, the model projects rapid epidemic declines: incidence and awareness targets are achieved by 2025 and sustained through 2030, while diagnoses fall to levels broadly consistent with national goals, though not in exact alignment. More gradual reductions in 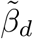 are sufficient to reach the 2030 incidence goal but leave diagnosis targets unmet. When both 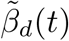 and *ρ*(*t*) are refitted after 2019, the model provides the closest match to observed post-2019 trends.

The concept of virtual epidemics extends the virtual patient framework to infectious disease modeling, generating multiple plausible epidemic trajectories that are consistent with observed surveillance data while accounting for parameter uncertainty and data limitations such as underreporting and reporting delays. Applying this framework to the U.S. HIV epidemic, the analysis reveals asymmetry in the achievability of the Ending the HIV Epidemic initiative’s targets: only 0.04% of virtual epidemics reached the 2025 incidence goal of 9,300 new infections, while 67.08% reached the 2030 goal of 3,000. Critically, no virtual epidemic met both targets simultaneously, suggesting that the short-term 2025 target is highly unlikely under realistic parameter uncertainty, even with favorable post-2019 reductions in treatment-related transmission. Nevertheless, the substantially higher probability of achieving the 2030 target suggests that continued investments in diagnosis, treatment, and prevention efforts may still support meaningful progress toward long-term HIV control. In summary, the virtual epidemic framework provides an important way to quantify prediction uncertainty and assess intervention feasibility, offering more subtle public health insight than single best-fit model projections.

Our epidemiological findings underscore that treatment-as-prevention is most effective when paired with earlier and broader diagnosis coverage. Multiscale models thus provide a rigorous framework for evaluating epidemic goals and identifying the intervention strategies most likely to accelerate progress toward long-term HIV control. This study demonstrates both the potential and the challenges of multiscale modeling for infectious diseases. Our approach combines structural and practical identifiability with sequential parameter estimation, providing a robust framework for modeling HIV and related epidemics.

## Data Availability

The datasets analyzed in this study were obtained from publicly available sources and from the Stanford HIV Drug Resistance Database with permission. The relevant data sources are cited in the manuscript. Additional information regarding the processed data is available from the corresponding author upon reasonable request.

## Appendix A Analysis of the Multiscale HIV Model

To understand the system’s dynamics, we first identify its equilibria and then analyze their stability properties. The equilibria of the between-host model are determined by the following system of equations:

**Table A1.**
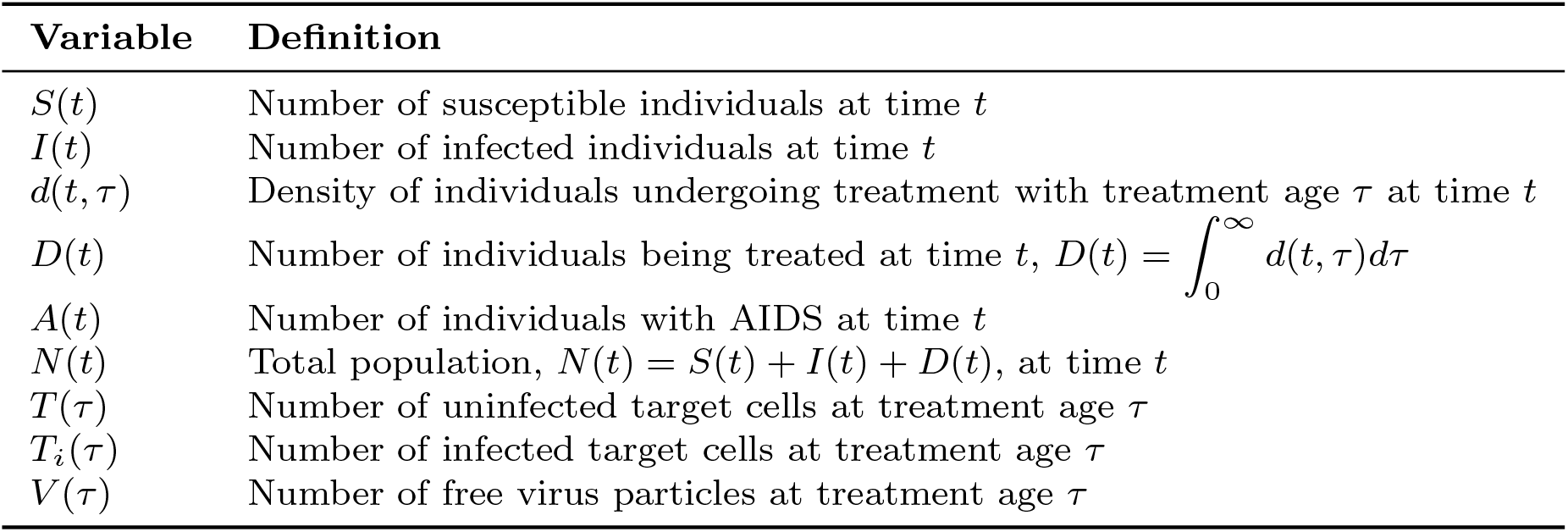
Definitions of the state variables of the multiscale HIV model (1).

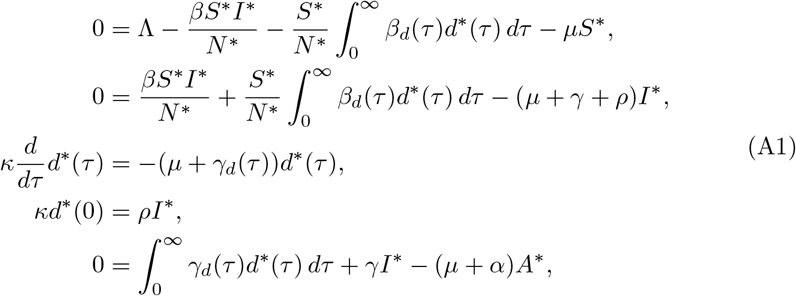

where 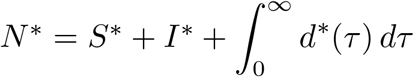. System (A1) has two equilibria: the disease-free equilibrium (DFE), representing the absence of disease in the population, and the endemic equilibrium (EE), representing the persistence of infection. We first analyze the DFE and its stability; then we derive the EE and determine its stability.

**Table A2.**
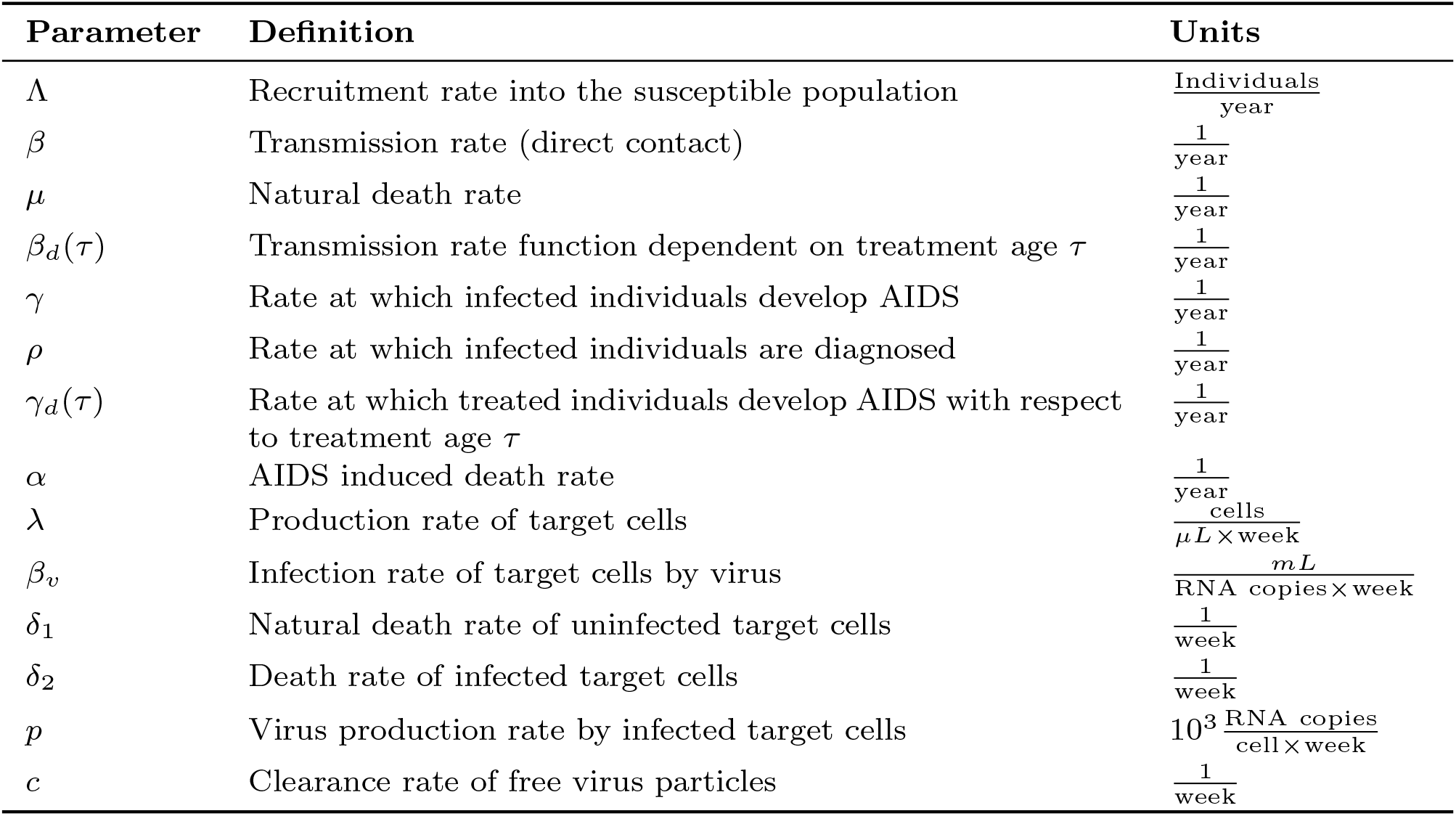
Model parameters in system (1), along with definitions and associated units. Population-level parameters use time units of years, whereas within-host parameters use time units of weeks.

### Disease-Free Equilibrium and Its Stability Analysis

The disease-free equilibrium (DFE) is represented by

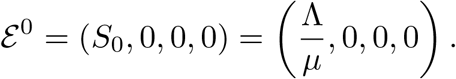

#### Theorem 3

*If R*_0_ *<* 1, *then the disease-free equilibrium E* ^0^ *is locally asymptotically stable, where the basic reproduction number R*_0_ *is given by*

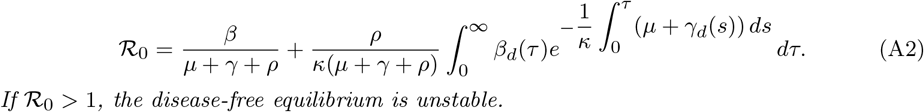

*If R*_0_ *>* 1, *the disease-free equilibrium is unstable*.

*Proof* To examine the stability of the disease-free equilibrium (DFE), we linearize the model (A1) around the point 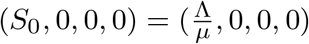. We introduce the following perturbations:

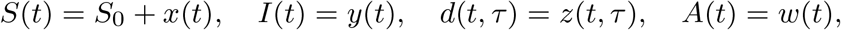

where 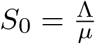 and the total population is

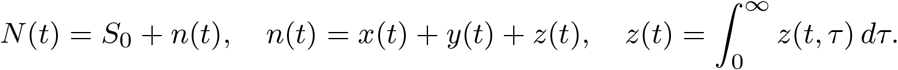

Expanding 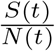 for small perturbations,

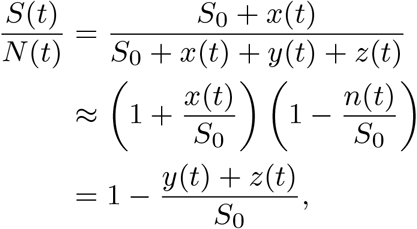

neglecting higher-order terms.

The linearized equation for *x*(*t*) can be obtained by the following.

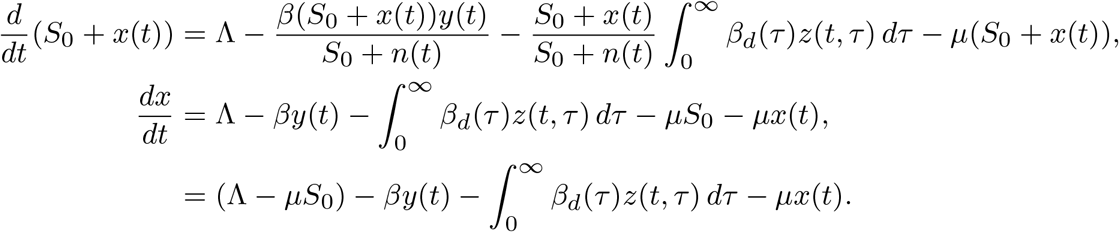

Since 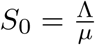, we have Λ − *µS*_0_ = 0, so the final linearized equation is

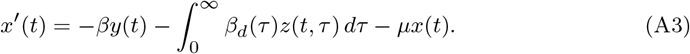

This equation, along with the linearized equations for *y*(*t*), *z*(*t, τ*), and *w*(*t*), forms the linearized system around the disease-free equilibrium.

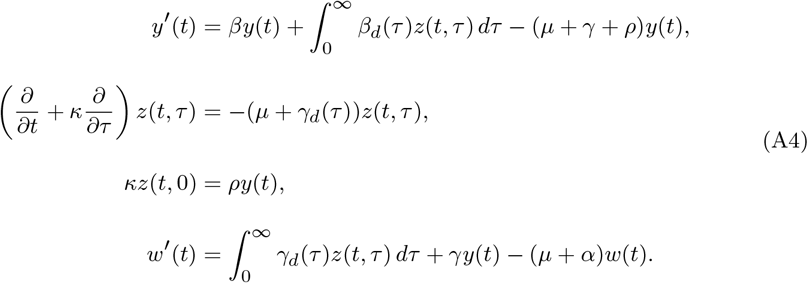

To evaluate the stability of the disease-free equilibrium, we seek solutions for the linearized system in the form of exponential functions.

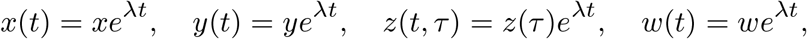

where *λ* ∈ C is the eigenvalue to be determined, with real part *a* = Re(*λ*) and imaginary part *b* = Im(*λ*). Substituting into the linearized system yields the eigenvalue problem:

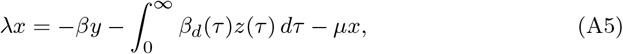

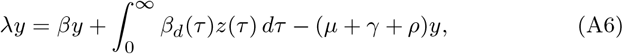

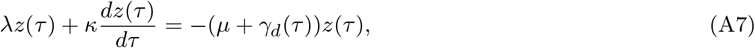

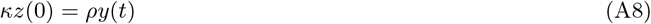

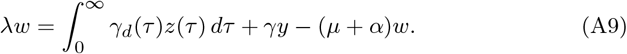

Solving the equation for *z*(*τ*) with initial condition, we obtain

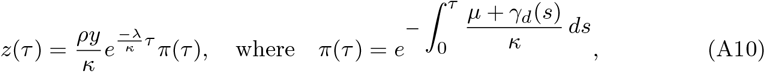

where *π*(*τ*) is the survival probability in the diagnosed (treated) compartment up to age *τ*. Substituting *z*(*τ*) into (A6) and assuming *y≠* 0, the characteristic equation reduces to

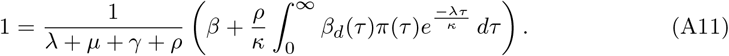

We define the right side of equation (A11) as *F* (*λ*), so that *F* (*λ*) = 1.

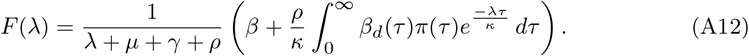

Note that, *F* (0) is the basic reproduction number *R*_0_ given by,

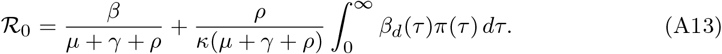

We proceed by contradiction: suppose there exists an eigenvalue *λ* = *a* + *bi* with *a* ≥ 0 when *R*_0_ *<* 1. The characteristic equation for the eigenvalue is *F* (*λ*) = 1. Let’s analyze |*F* (*λ*)|.

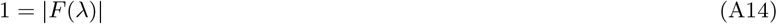

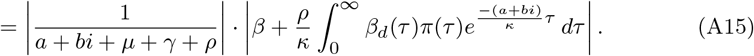

To analyze |*F* (*λ*)|, we employ the properties of complex modulus, particularly utilizing the fact that for a complex number 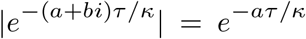. This key property allows us to simplify the integral term in our analysis.

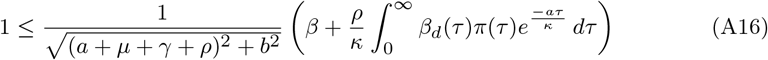

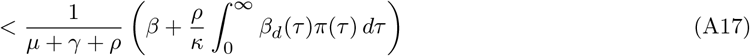

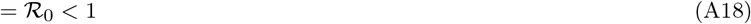

This chain of inequalities leads to a contradiction. Therefore, all eigenvalues satisfy Re(*λ*) *<* 0 when *R*_0_ *<* 1, so the DFE is locally asymptotically stable. On the other hand, if *R*_0_ *>* 1, then *F* (0) = *R*_0_ *>* 1 and lim_*λ*→∞_ *F* (*λ*) = 0. By continuity, there exists *λ*\* *>* 0 such that *F* (*λ*\*) = 1, which implies the existence of a positive real eigenvalue. Hence, the DFE, *E*_0_ is unstable when *R*_0_ *>* 1. □

### A.0.1 Endemic equilibrium and its stability analysis

To obtain the endemic equilibrium, *ℰ*\* = (*S*\*, *I*\*, *d*\*(*τ*), *A*\*), we first solve the differential equation for *d*\*(*τ*) from (A1), yielding

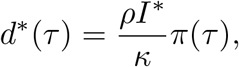

where

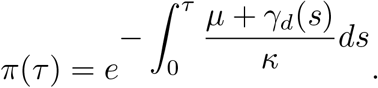

Next, adding the first two equations in (A1), we obtain 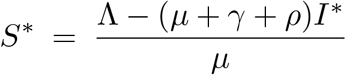.

Integrating the third equation (A1) with respect to *τ* we get, and using the boundary condition lim_*τ*→∞_ *d*\*(*τ*) = 0, we have

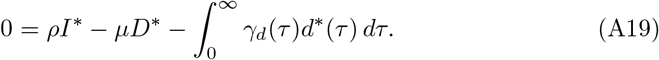

Adding the first two equations in (A1) and (A19), we obtain

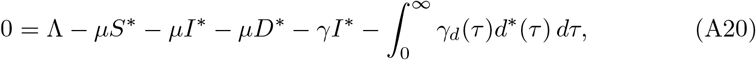

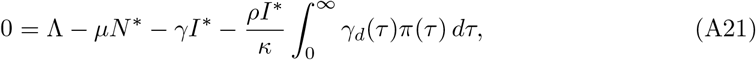

where

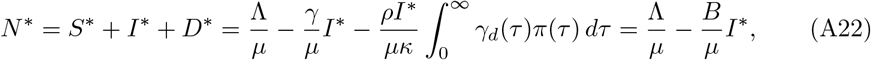

with 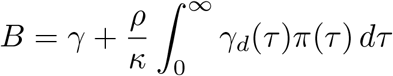.

Since *I*\* */*= 0 at the endemic equilibrium, the second equilibrium equation in (A1), gives

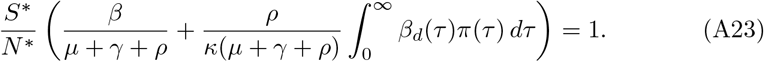

Substituting *S*\* and *N*\* into (A23), we obtain the following expression, which we define as *G*(*I*\*):

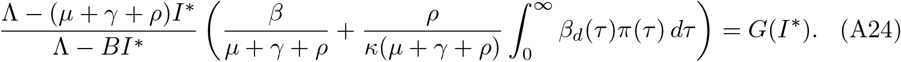

First, we show that *B* − (*µ* + *γ* + *ρ*) *<* 0. Note that,

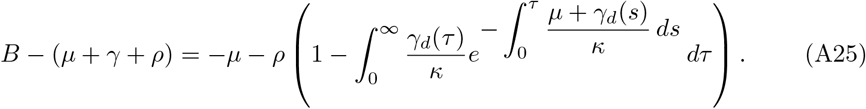

Next, we will show that

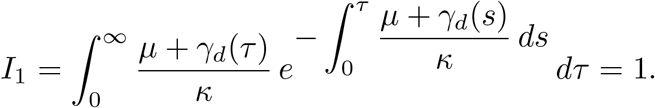

Note that

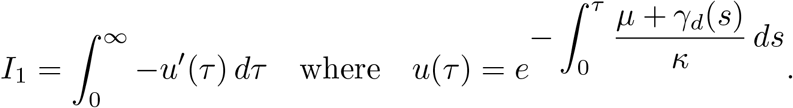

Clearly *I*_1_ = 1. Therefore,

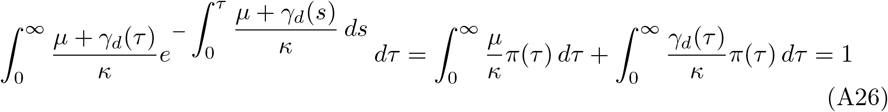

We conclude that 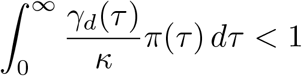, which proves that *B* − (*µ* + *γ* + *ρ*) *<* 0. We rewrite the equation (A24), and we get

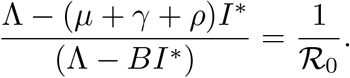

Solving for *I*\*, we get

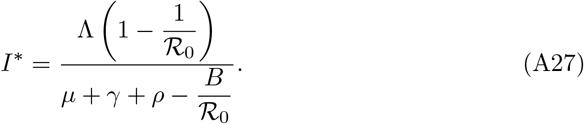

Note that since *R*_0_ *>* 1, both the numerator and the denominator of this expression are positive, which ensures that *I*\* is well defined and strictly positive. Once *I*\* is computed, the equilibrium values of *N*\*, *S*\*, and *d*\*(*τ*) can be determined. These quantities are given by the following equations.

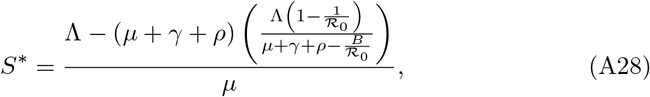

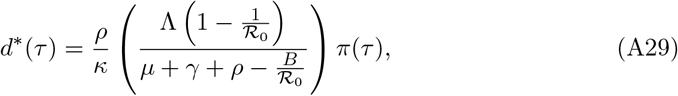

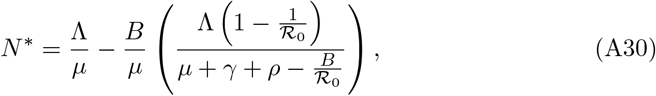

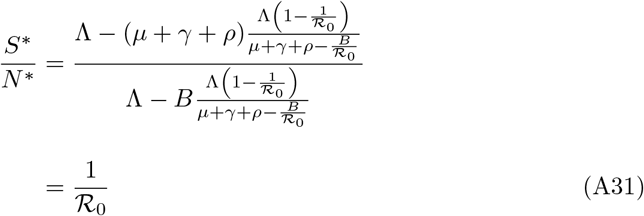

Therefore, we show that *S*\**/N*\* = 1*/R*_0_.

#### Theorem 4

*If* 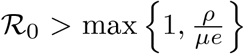, *then the endemic equilibrium (EE) is locally asymptotically stable*.

*Proof* To study the stability of the endemic equilibrium, we linearize the model around the endemic equilibrium (*S*\*, *I*\*, *d*\*(*τ*), *A*\*). We introduce the following perturbation variables.

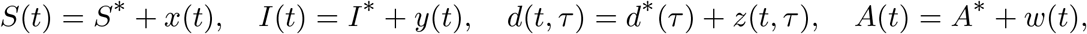

where 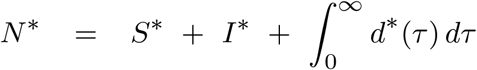, and *N* (*t*) = *N* * + *n*(*t*) with 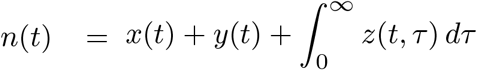

The system with perturbations satisfies the following differential equations.

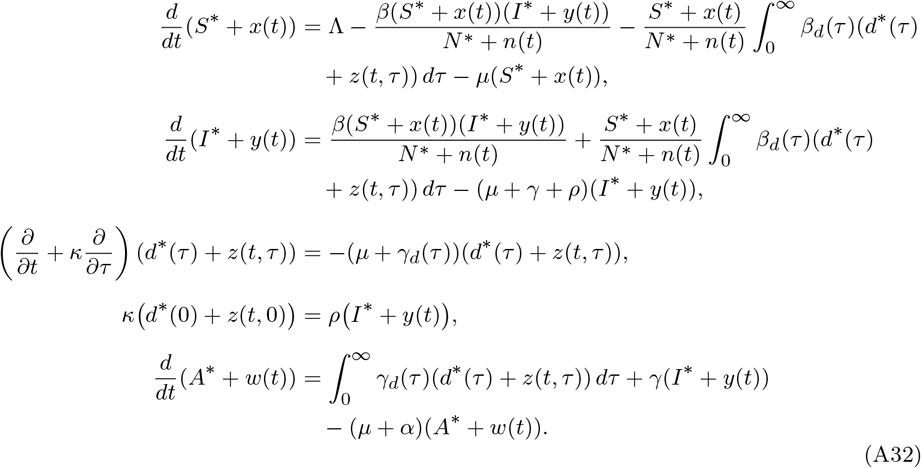

We write the fraction 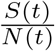 as:

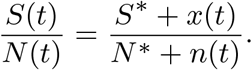

To approximate 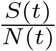, using a first-order Taylor expansion around the equilibrium values, assuming that *x*(*t*) and *n*(*t*) are small perturbations. First, we express the denominator as,

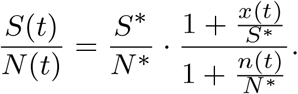

Using a first-order approximation for 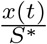 and 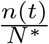

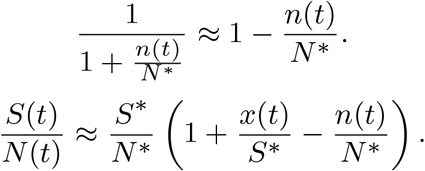

Therefore, the approximation for 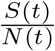 is

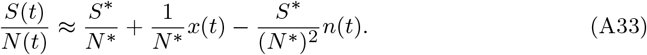

The approximation for 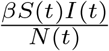 is updated.

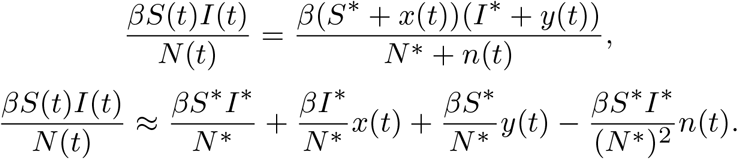

We linearize the system at the endemic equilibrium.

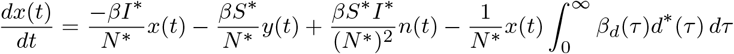

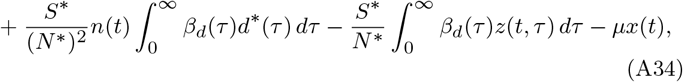

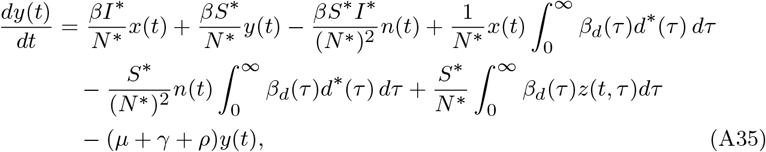

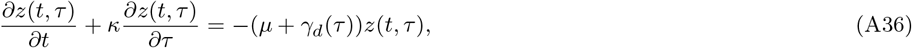

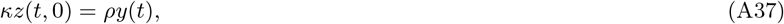

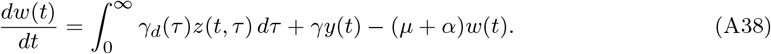

To assess the local stability of the endemic equilibrium, we analyze the solutions of the linearized system. These solutions are expected to be exponential in form.

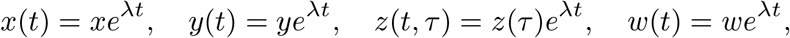

where *x, y, w* are constants, *z*(*τ*) is a function of *τ*, and *λ* is the eigenvalue. Substituting these expressions into the linearized system and dividing through by *e*^*λt*^ throughout, we obtain the following linear eigenvalue problem for *x, y, z*(*τ*), *w*, and the eigenvalue *λ*.

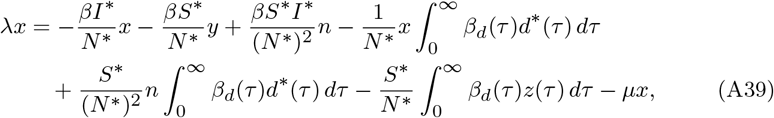

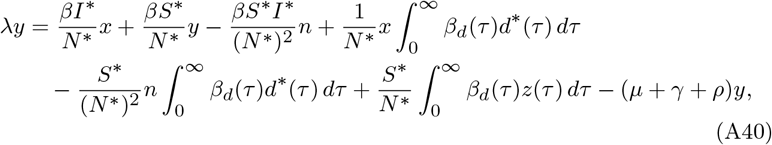

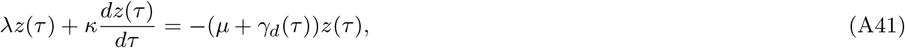

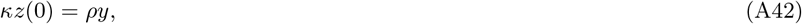

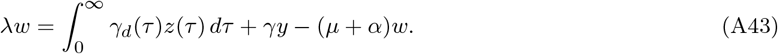

This system of equations forms an eigenvalue problem. The eigenvalues, *λ* that satisfy this system determine the stability of the endemic equilibrium. We first solve the equation (A41) for *z*(*τ*).

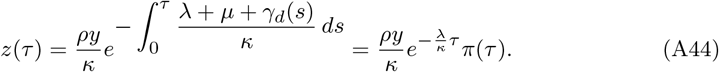

Adding the first two equations (A39) and (A40), we have

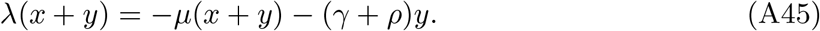

Next, integrating the third equation (A41), we obtain

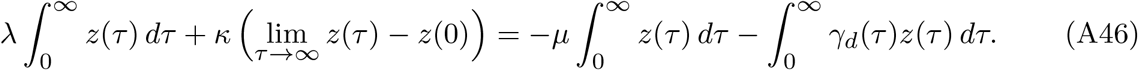

Using the boundary condition *κz*(0) = *ρy*, and observing that lim*τ* →∞ *z*(*τ*) = 0 due to the natural decay of *z*(*τ*), this simplifies to,

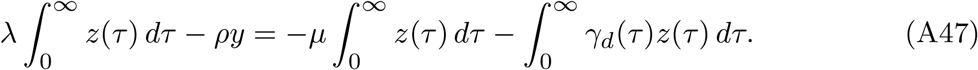

Combining the equations for *x* + *y* and 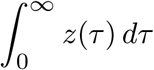, we obtain

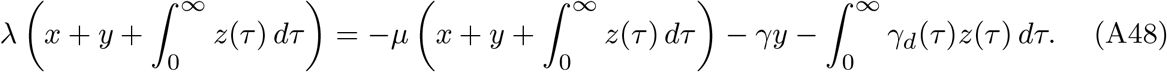

Since 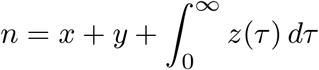, and 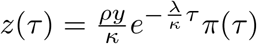

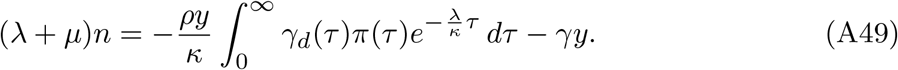

We add the first two equations (A39) and (A40) and solve for *x*

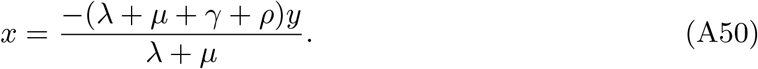

We substitute equation (A50) into equation (A40) and denote the following normalized quantities as follows:

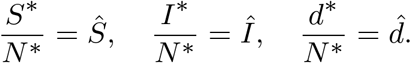

This leads to the following expression.

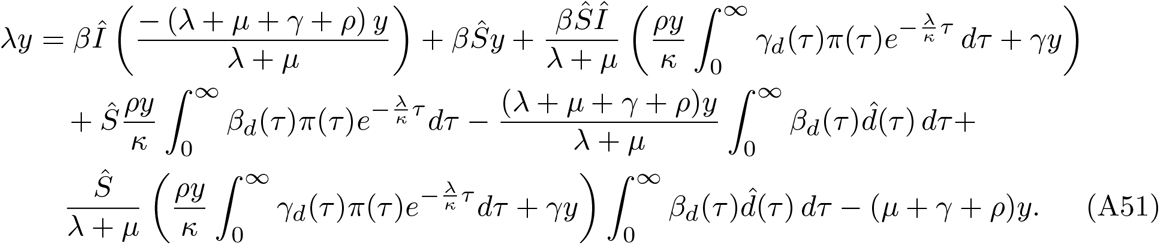

Since *y≠* 0, simplifying *y*, we obtain the following characteristic equation.

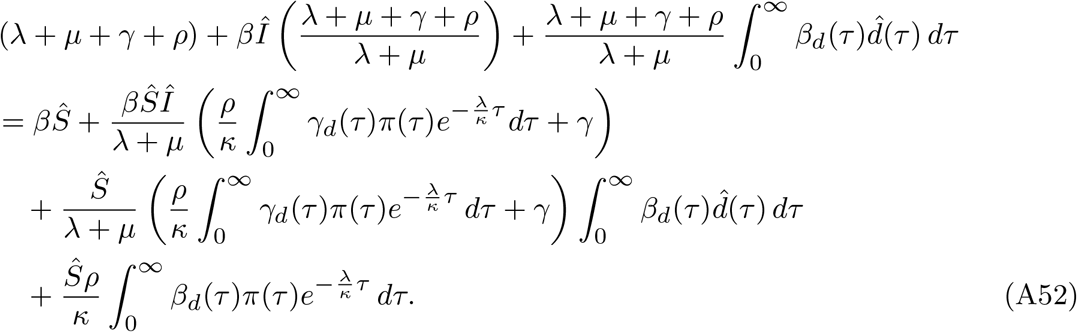

After applying the condition 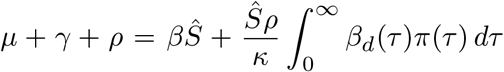 from the second and third equations (A1), we rewrite the characteristic equation as:

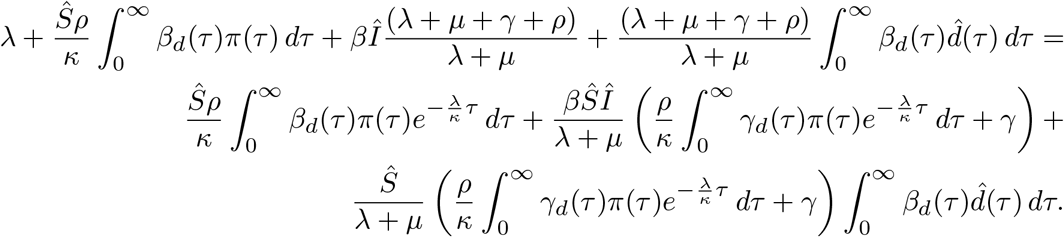

Rearranging the characteristic equation, we obtain

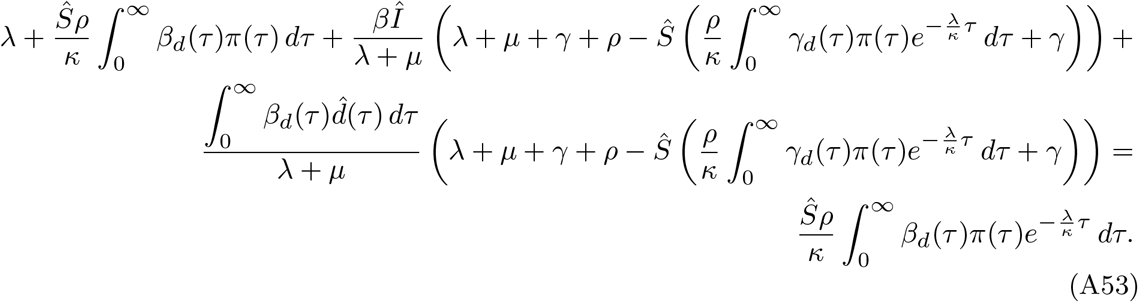

To analyze the stability, we separate the characteristic equation into two parts. We denote the left-hand side of (A53) as *H*(*λ*) and the right-hand side as *K*(*λ*), rewriting the equation as

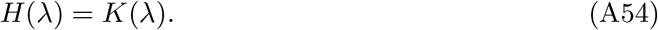

We begin by examining the right-hand side of the characteristic equation (A53), denoted as

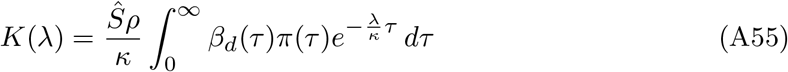

Assuming *λ* = *a* + *bi* where *a >* 0, we can bound |*K*(*λ*)| as follows

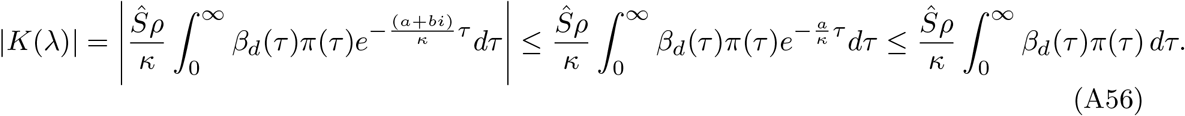

Next, we analyze the left-hand side of equation (A53), denoted as *H*(*λ*)

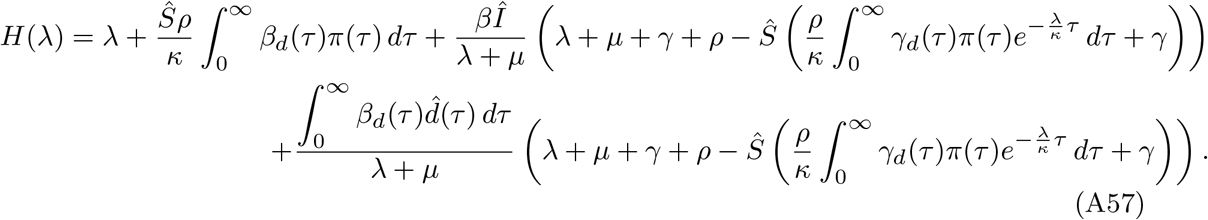

We substitute *λ* = *a* + *bi* for *H*(*λ*) and we get,

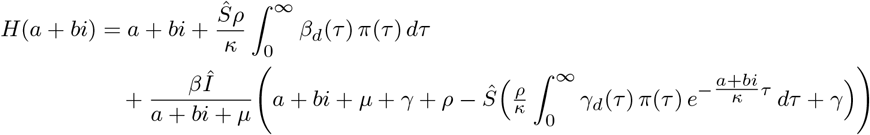

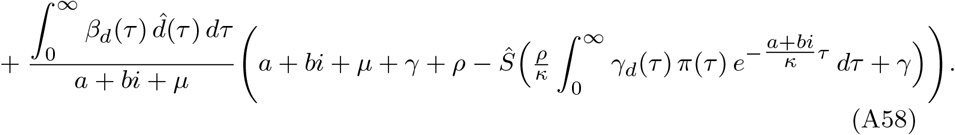

To simplify this expression, we focus on the following term, and we separate the real and imaginary parts of the integral.

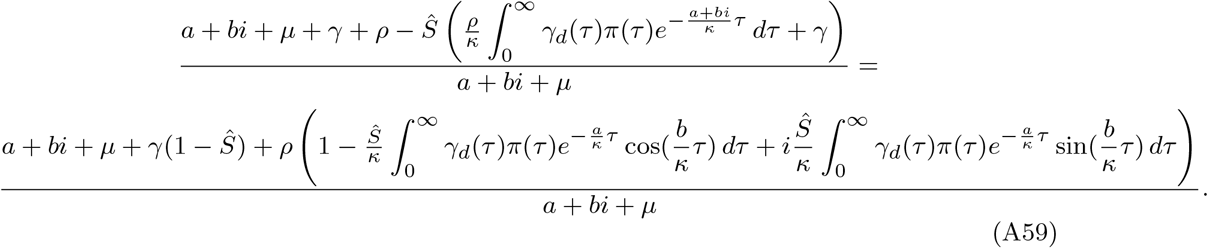

For simplicity, we define the following terms.

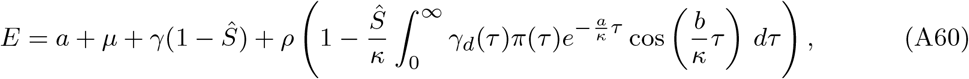

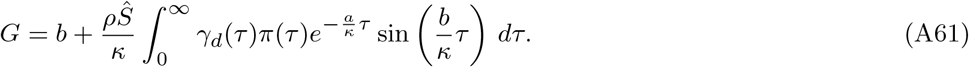

Thus, the fraction simplifies to

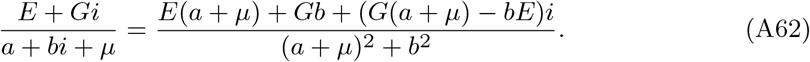

Taking the magnitude of equation *H*(*λ*) = *K*(*λ*), we obtain:

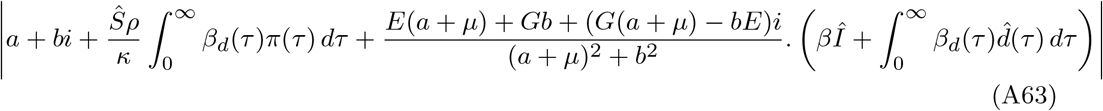

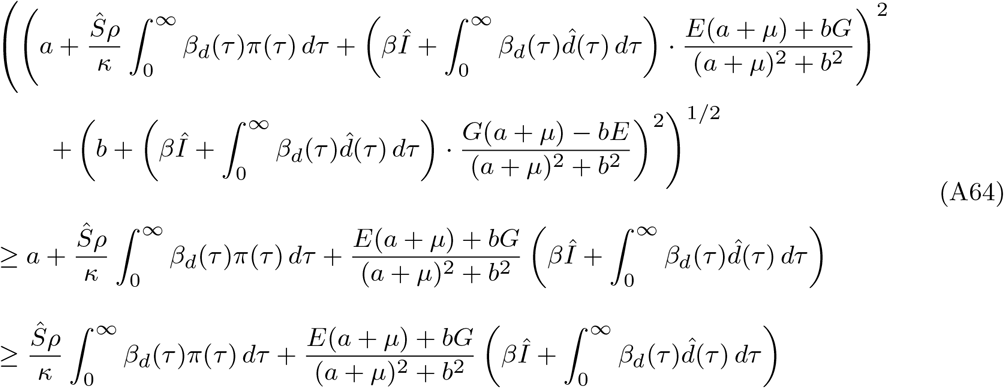

We now simplify this inequality by considering two key conditions: First, consider the positivity condition 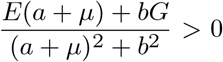. Second, the assumption Ŝ *<* 1 holds. From the definition of E, we can isolate the term *γ*(1 − Ŝ) *>* 0. Thus, we can write

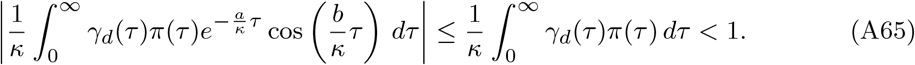

We conclude that *E* ≥ 0 and since the imaginary part of *a* + *bi* can be taken as positive, we also have *b* ≥ 0.

To analyze the term *G* we can proceed as follows. For the integral part of *G*, we can apply the inequality sin(*x*) ≤ *x* for *x* ≥ 0.

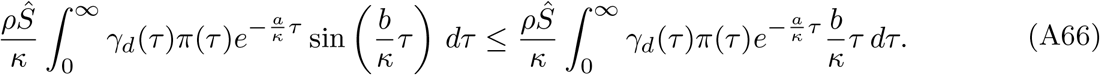

Recall the definition of *π*(*τ*)

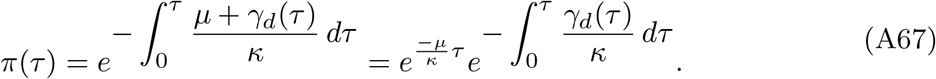

We can further simplify the integral.

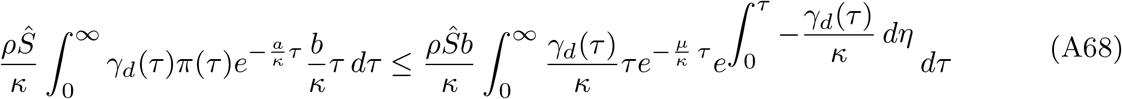

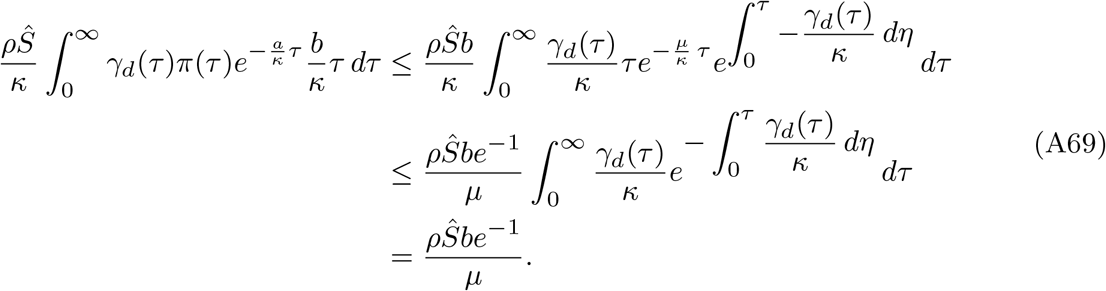

Therefore, we can establish a lower bound for *G*,

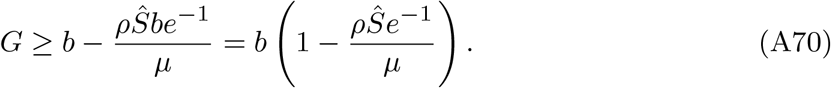

For the endemic equilibrium (EE) to be locally asymptotically stable, we require *G >* 0 for all *b* ∈ R^+^. A sufficient condition for this inequality to hold is 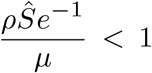. Substituting 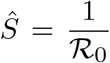 from (A31), this condition becomes 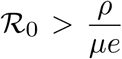 which establishes 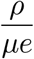 as a critical threshold for the local asymptotic stability of the endemic equilibrium. Furthermore, under this condition we obtain the bounds

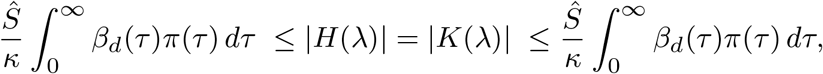

which leads to a contradiction. Therefore, the characteristic equation *H*(*λ*) = *K*(*λ*) does not admit roots with nonnegative real parts when 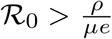. □

## Appendix B HIV Data at Multiple Epidemiological Scales

## Appendix C Structural identifiability – input-output equation for multiscale HIV model

The following expanded expression corresponds to the second input-output equation derived from the population-level subsystem after eliminating unobservable variables. The equation relates the observable outputs *y*_1_(*t*), *y*_2_(*t*), and *y*_3_(*t*) to the model parameters and is used in the structural identifiability analysis of the multiscale HIV model.

~~~
In[2] : = (*Definitions: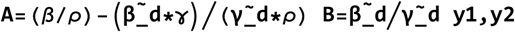
y3=state variables y_1_(t),y_2_(t),y_3_(t) yp1,yp2,yp3=first derivatives:y_1_*′*(t),
y_2_*′*(t),y_3_*′*(t) ypp1,ypp2,ypp3=second derivatives:y_1_*″*(t),
y_2_*″*(t),y_3_*″*(t) Lam=*λ* (a parameter),mu=*μ* (another parameter)*)
Expand[(2 (Lam - y1) (A y3 + B y2) (A yp3 + B yp2) - yp1 (A y3 + B y2) ^ 2 + yp1 y2 (A y3 + B y2) +
    y1 yp2 (A y3 + B y2) + y1 y2 (A yp3 + B yp2) - Lam (yp1 (A y3 + B y2) + y1 (A yp3 + B yp2)))
 (yp1 (A y3 + B y2) - y1 (A yp3 + B yp2)) - (Lam (A y3 + B y2) ^ 2 - y1 (A y3 + B y2) ^ 2 +
 y1 y2 (A y3 + B y2) - Lam y1 (A y3 + B y2)) (ypp1 (A y3 + B y2) - y1 (A ypp3 + B ypp2)) -
 (Lam - y2) (yp1 (A y3 + B y2) - y1 (A yp3 + B yp2)) ^ 2 +
 mu (Lam (A y3 + B y2) ^ 2 - y1 (A y3 + B y2) ^ 2 + y1 y2 (A y3 + B y2) - Lam y1 (A y3 + B y2))
 (yp1 (A y3 + B y2) - y1 (A yp3 + B yp2))]
Out[2] = −B^2^ Lam mu y1 y2^2^ yp1 + B^3^ Lam mu y2^3^ yp1 + B^2^ mu y1 y2^3^ yp1 - B^3^ mu y1 y2^3^ yp1 - 2 A B Lam mu y1 y2 y3 yp1 +
3 A B^2^ Lam mu y2^2^ y3 yp1 + 2 A B mu y1 y2^2^ y3 yp1 - 3 A B^2^ mu y1 y2^2^ y3 yp1 - A^2^ Lam mu y1 y3^2^ yp1 +
3 A^2^ B Lam mu y2 y3^2^ yp1 + A^2^ mu y1 y2 y3^2^ yp1 - 3 A^2^ B mu y1 y2 y3^2^ yp1 + A^3^ Lam mu y3^3^ yp1 -
A^3^ mu y1 y3^3^ yp1 - 2 B^2^ Lam y2^2^ yp1^2^ + 2 B^2^ y2^3^ yp1^2^ - B^3^ y2^3^ yp1^2^ -  4 A B Lam y2 y3 yp1^2^ +
4 A B y2^2^ y3 yp1^2^ - 3 A B^2^ y2^2^ y3 yp1^2^ - 2 A^2^ Lam y3^2^ yp1^2^ + 2 A^2^ y2 y3^2^ yp1^2^ - 3 A^2^ B y2 y3^2^ yp1^2^ -
A^3^ y3^3^ yp1^2^ + B^2^ Lam mu y1^2^ y2 yp2 - B^3^ Lam mu y1 y2^2^ yp2 - B^2^ mu y1^2^ y2^2^ yp2 + B^3^ mu y1^2^ y2^2^ yp2 +
A B Lam mu y1^2^ y3 yp2 - 2 A B^2^ Lam mu y1 y2 y3 yp2 - A B mu y1^2^ y2 y3 yp2 + 2 A B^2^ mu y1^2^ y2 y3 yp2 -
A^2^ B Lam mu y1 y3^2^ yp2 + A^2^ B mu y1^2^ y3^2^ yp2 + 2 B^2^ Lam y1 y2 yp1 yp2 + 2 B^3^ Lam y2^2^ yp1 yp2 -
B^2^ y1 y2^2^ yp1 yp2 - B^3^ y1 y2^2^ yp1 yp2 + 2 A B Lam y1 y3 yp1 yp2 + 4 A B^2^ Lam y2 y3 yp1 yp2 - 2 A B^2^ y1 y2 y3 yp1 yp2 +
2A^2^ B Lam y3^2^ yp1 yp2 + A^2^ y1 y3^2^ yp1 yp2 - A^2^ B y1 y3^2^ yp1 yp2 -
2 B^3^ Lam y1 y2 yp2^2^ - B^2^ y1^2^ y2 yp2^2^ + 2 B^3^ y1^2^ y2 yp2^2^ - 2 A B^2^ Lam y1 y3 yp2^2^ - A B y1^2^ y3 yp2^2^ +
2 A B^2^ y1^2^ y3 yp2^2^ + A B Lam mu y1^2^ y2 yp3 - A B^2^ Lam mu y1 y2^2^ yp3 - A B mu y1^2^ y2^2^ yp3 +
A B^2^ mu y1^2^ y2^2^ yp3 + A^2^ Lam mu y1^2^ y3 yp3 - 2 A^2^ B Lam mu y1 y2 y3 yp3 - A^2^ mu y1^2^ y2 y3 yp3 +
2 A^2^ B mu y1^2^ y2 y3 yp3 - A^3^ Lam mu y1 y3^2^ yp3 + A^3^ mu y1^2^ y3^2^ yp3 + 2 A B Lam y1 y2 yp1 yp3 +
2 A B^2^ Lam y2^2^ yp1 yp3 - 2 A B y1 y2^2^ yp1 yp3 - A B^2^ y1 y2^2^ yp1 yp3 + 2 A^2^ Lam y1 y3 yp1 yp3 +
4 A^2^ B Lam y2 y3 yp1 yp3 - 2 A^2^ y1 y2 y3 yp1 yp3 - 2 A^2^ B y1 y2 y3 yp1 yp3 + 2 A^3^ Lam y3^2^ yp1 yp3 -
A^3^ y1 y3^2^ yp1 yp3 - 4 A B^2^ Lam y1 y2 yp2 yp3 - A B y1^2^ y2 yp2 yp3 + 4 A B^2^ y1^2^ y2 yp2 yp3 -
4 A^2^ B Lam y1 y3 yp2 yp3 - A^2^ y1^2^ y3 yp2 yp3 + 4 A^2^ B y1^2^ y3 yp2 yp3 - 2 A^2^ B Lam y1 y2 yp3^2^ +
2 A^2^ B y1^2^ y2 yp3^2^ - 2 A^3^ Lam y1 y3 yp3^2^ + 2 A^3^ y1^2^ y3 yp3^2^ + B^2^ Lam y1 y2^2^ ypp1 - B^3^ Lam y2^3^ ypp1 -
B^2^ y1 y2^3^ ypp1 + B^3^ y1 y2^3^ ypp1 + 2 A B Lam y1 y2 y3 ypp1 - 3 A B^2^ Lam y2^2^ y3 ypp1 -
2 A B y1 y2^2^ y3 ypp1 + 3 A B^2^ y1 y2^2^ y3 ypp1 + A^2^ Lam y1 y3^2^ ypp1 - 3 A^2^ B Lam y2 y3^2^ ypp1 -
A^2^ y1 y2 y3^2^ ypp1 + 3 A^2^ B y1 y2 y3^2^ ypp1 - A^3^ Lam y3^3^ ypp1 + A^3^ y1 y3^3^ ypp1 - B^2^ Lam y1^2^ y2 ypp2 +
B^3^ Lam y1 y2^2^ ypp2 + B^2^ y1^2^ y2^2^ ypp2 - B^3^ y1^2^ y2^2^ ypp2 - A B Lam y1^2^ y3 ypp2 + 2 A B^2^ Lam y1 y2 y3 ypp2 +
A B y1^2^ y2 y3 ypp2 - 2 A B^2^ y1^2^ y2 y3 ypp2 + A^2^ B Lam y1 y3^2^ ypp2 - A^2^ B y1^2^ y3^2^ ypp2 -
A B Lam y1^2^ y2 ypp3 + A B^2^ Lam y1 y2^2^ ypp3 + A B y1^2^ y2^2^ ypp3 - A B^2^ y1^2^ y2^2^ ypp3 - A^2^ Lam y1^2^ y3 ypp3 +
2 A^2^ B Lam y1 y2 y3 ypp3 + A^2^ y1^2^ y2 y3 ypp3 - 2 A^2^ B y1^2^ y2 y3 ypp3 + A^3^ Lam y1 y3^2^ ypp3 - A^3^ y1^2^ y3^2^ ypp3
~~~

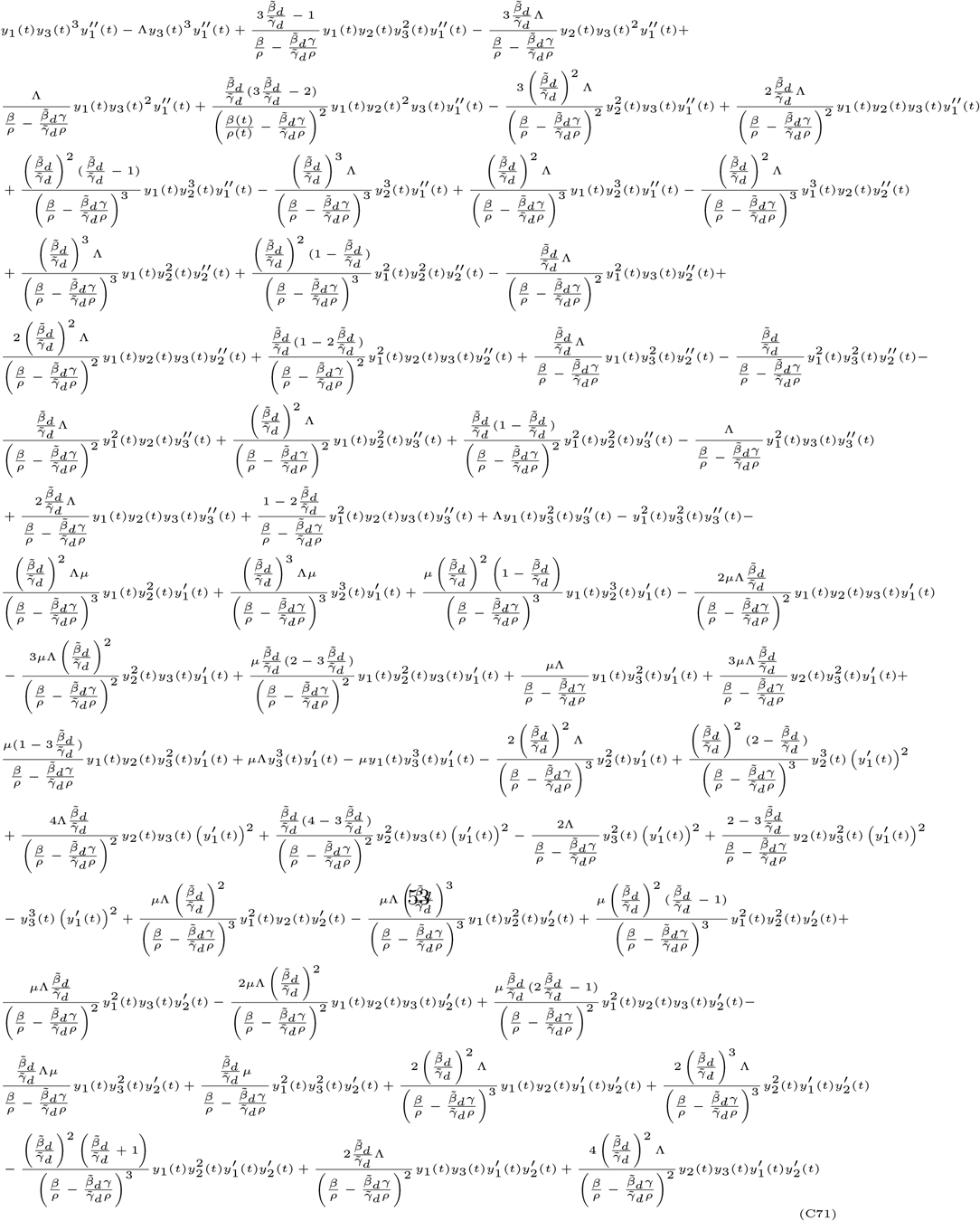

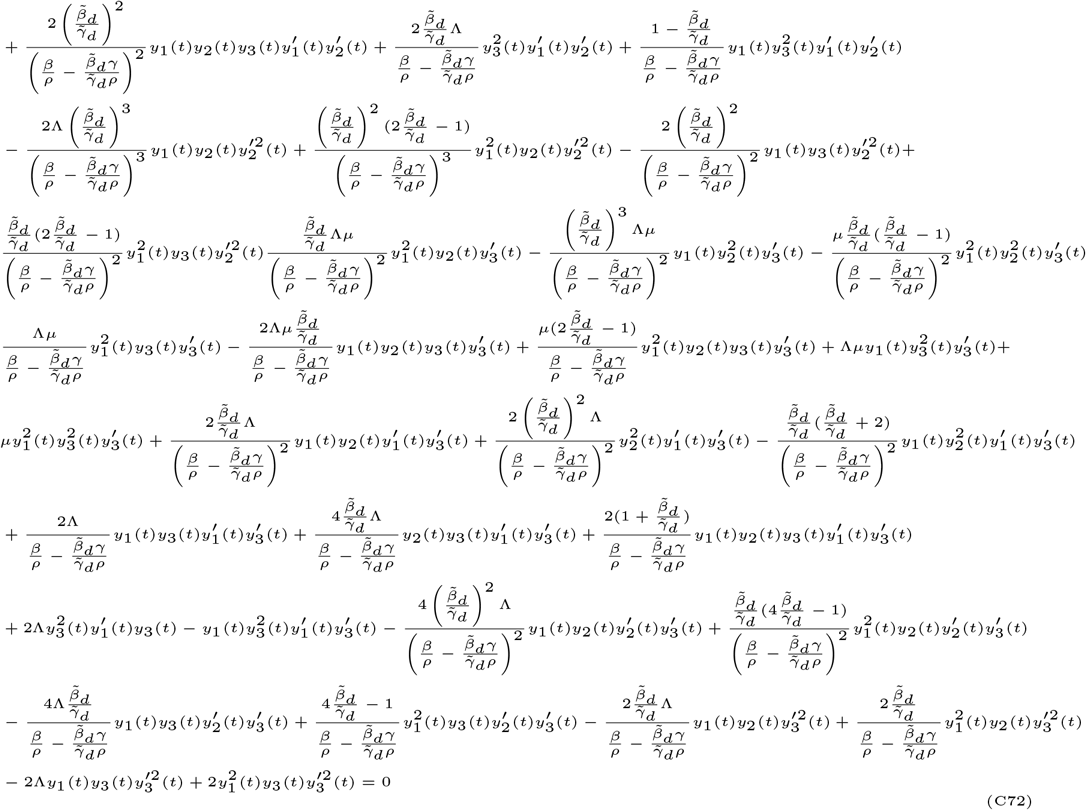

**Table B3.** Estimated annual HIV cases (in thousands) in the United States, based on CDC data [38]. In the multiscale HIV model (1), the observed HIV incidence is given by 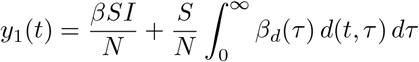.

| Year | HIV Incidents (1000s) | Year | HIV Incidents (1000s) |
| --- | --- | --- | --- |
| 1981 | 20.77 | 2007 | 53.70 |
| 1982 | 65.39 | 2008 | 42.21 |
| 1983 | 65.13 | 2009 | 41.10 |
| 1984 | 131.07 | 2010 | 40.28 |
| 1985 | 131.37 | 2011 | 38.89 |
| 1986 | 85.37 | 2012 | 38.90 |
| 1990 | 85.15 | 2013 | 38.64 |
| 1991 | 49.53 | 2014 | 38.66 |
| 1996 | 49.89 | 2015 | 38.95 |
| 1997 | 58.60 | 2016 | 38.68 |
| 1999 | 59.19 | 2017 | 37.85 |
| 2000 | 56.40 | 2018 | 37.30 |
| 2006 | 56.49 | 2019 | 36.19 |

**Table B4.** Annual AIDS classification cases in the United States (in thousands), as reported by the CDC [36]. According to the multiscale HIV model (1), the observation of AIDS classifications is given by 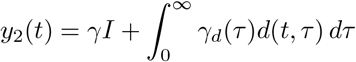.

| Years Since Classification | AIDS Cases (1000s) | Years Since Classification | AIDS Cases (1000s) |
| --- | --- | --- | --- |
| 1985 | 11.81 | 2003 | 36.99 |
| 1986 | 18.81 | 2004 | 35.90 |
| 1987 | 27.93 | 2005 | 33.96 |
| 1988 | 34.43 | 2006 | 32.62 |
| 1989 | 41.59 | 2007 | 31.82 |
| 1990 | 47.39 | 2008 | 31.23 |
| 1991 | 57.22 | 2009 | 30.06 |
| 1992 | 69.71 | 2010 | 27.33 |
| 1993 | 73.97 | 2011 | 25.56 |
| 1994 | 67.47 | 2012 | 24.61 |
| 1995 | 64.35 | 2013 | 23.57 |
| 1996 | 56.50 | 2014 | 19.24 |
| 1997 | 45.95 | 2015 | 18.54 |
| 1998 | 39.34 | 2016 | 18.28 |
| 1999 | 37.73 | 2017 | 17.73 |
| 2000 | 38.05 | 2018 | 17.11 |
| 2001 | 36.62 | 2019 | 16.41 |
| 2002 | 36.43 |  |  |

**Fig. B1.**
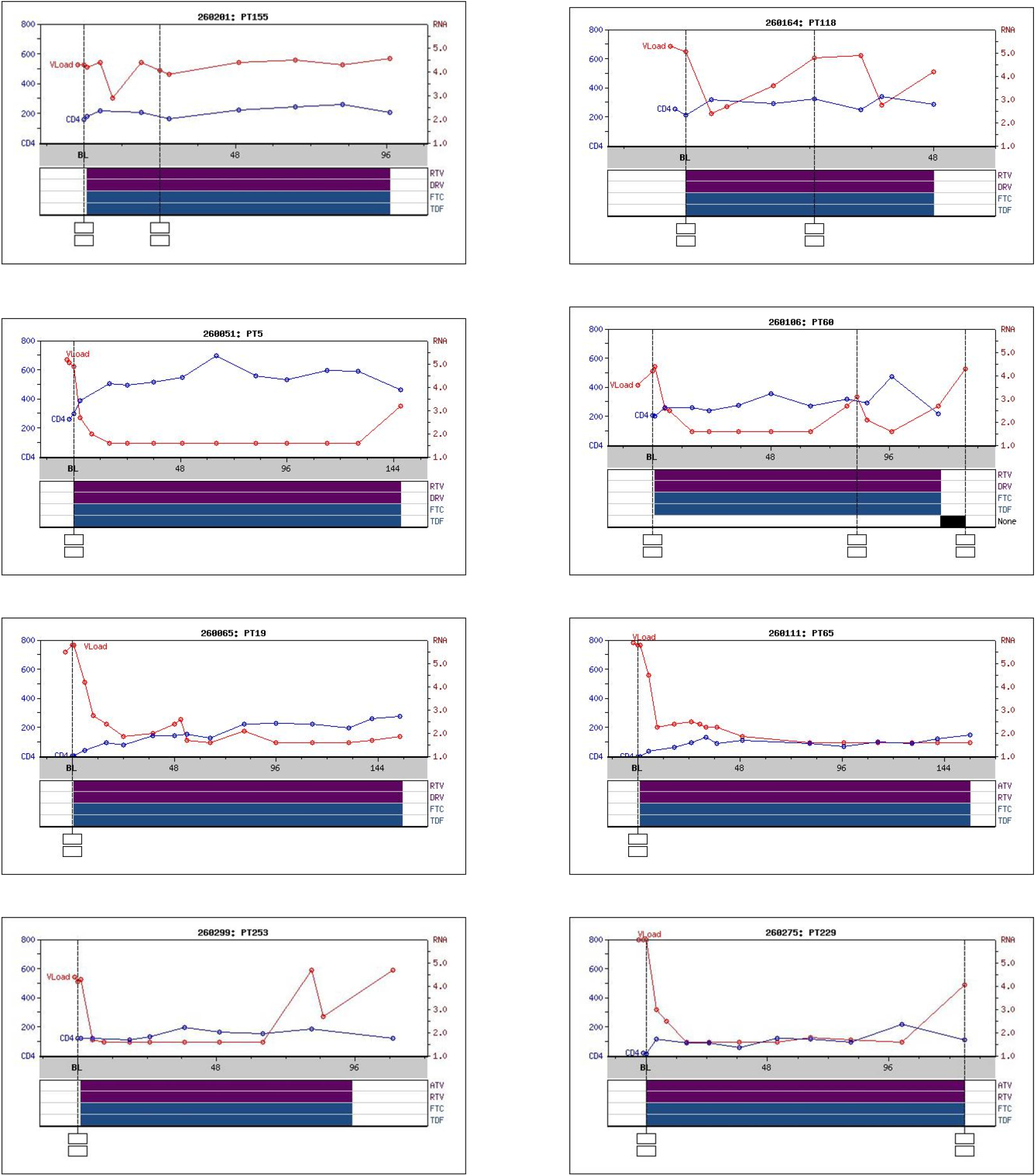
Trends in CD4^+^ cell count (blue line) and viral load (red line) over time for 8 representative patients from a cohort of 80 in the ACTG A5257 trial [39]. The gray shaded area indicates the treatment phase with antiretroviral drugs: NRTIs (FTC, TDF) in blue and PIs (RTV, DRV, ATV) in purple. Treatment duration varies by patient, with time measured in weeks.

**Table B5.**
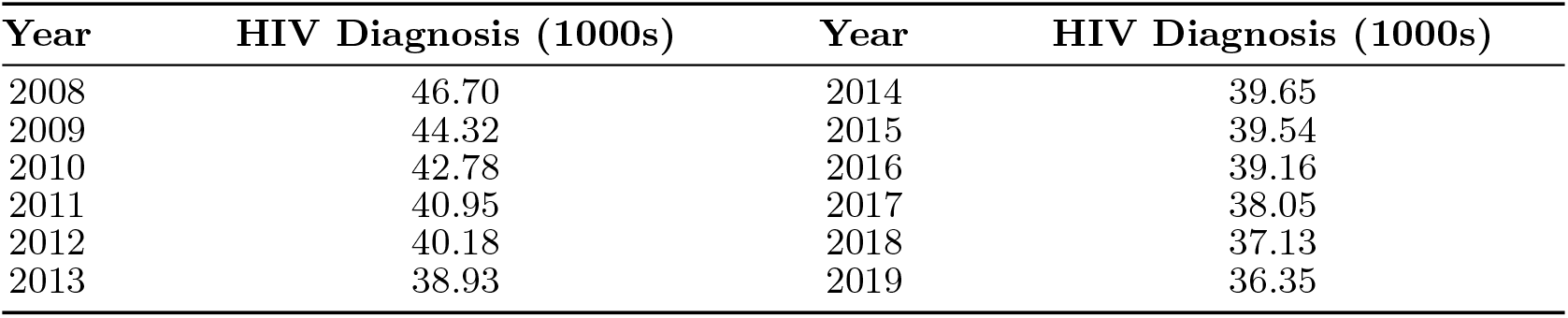
Annual HIV diagnosis cases in the United States (in thousands), as reported by the CDC [36]. In the multiscale model (1), the observed yearly diagnoses are represented by *y*_3_(*t*) = *ρI*.

| Year | HIV Diagnosis (1000s) | Year | HIV Diagnosis (1000s) |
| --- | --- | --- | --- |
| 2008 | 46.70 | 2014 | 39.65 |
| 2009 | 44.32 | 2015 | 39.54 |
| 2010 | 42.78 | 2016 | 39.16 |
| 2011 | 40.95 | 2017 | 38.05 |
| 2012 | 40.18 | 2018 | 37.13 |
| 2013 | 38.93 | 2019 | 36.35 |

## Appendix D Finite difference method

We construct a numerical method and implement a computational model for the system. The numerical approach is based on a finite difference scheme, which discretizes both the time and the treatment age variables. We first discretize the domain of the system. The independent variables in the model are time *t* and treatment age *τ*, both of which are approximated using discrete steps. The treatment age variable, *τ*, is discretized in the interval [0, *τ*_*f*_] with a step size Δ*τ*, such that

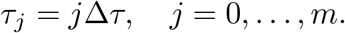

where *m*Δ*τ* = *τ*_*f*_.

Similarly, the variable *t* is discretized in the interval [0, *t*_*f*_] with a step size Δ*t*, so that

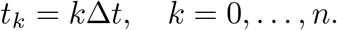

where *n*Δ*t* = *t*_*f*_.

The state variables are approximated at these discrete points. The susceptible population at time *t*_*k*_ is denoted by *S*^*k*^ ≈ *S*(*t*_*k*_), representing the number of individuals susceptible to infection. The infected population at time *t*_*k*_ is denoted by *I*^*k*^ ≈ *I*(*t*_*k*_), representing individuals who are currently infected and capable of spreading the disease. The treated population is indicated by 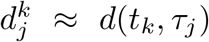, which represents individuals at time *t*_*k*_ who have received treatment for a period of *τ*_*j*_. The partial derivatives are approximated using a forward difference for time and a backward difference for treatment age.

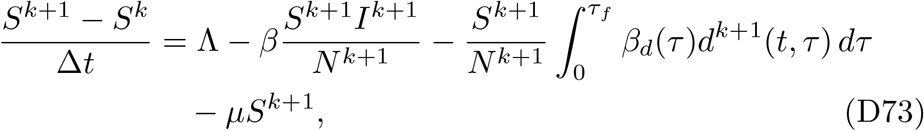

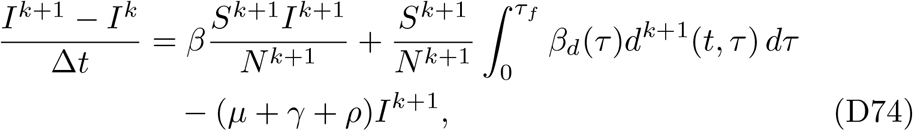

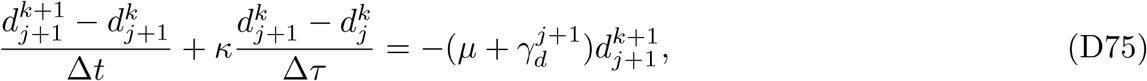

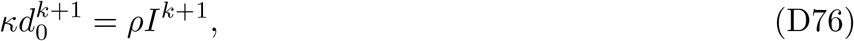

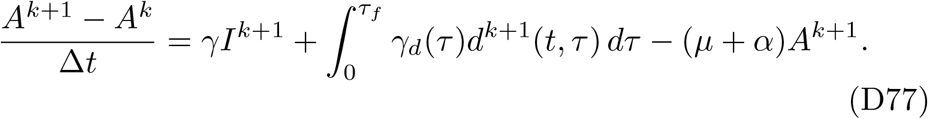

Since this system is non-linear due to the infection terms and their interaction with delayed compartments, we applied a Picard iteration to linearize it. To do this, we rewrite the system by replacing the non-linear terms with values from the previous iteration or time step. Applying Picard iteration, we obtain the linearized system as follows:

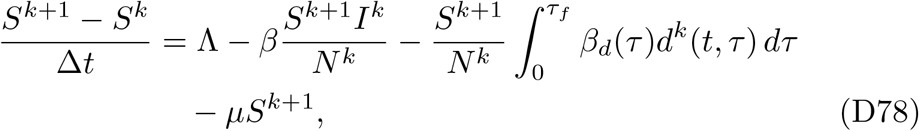

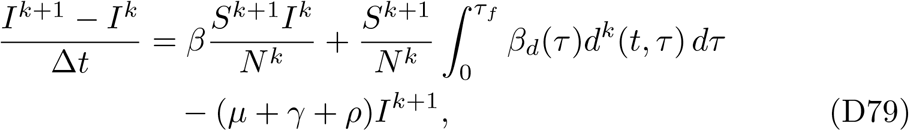

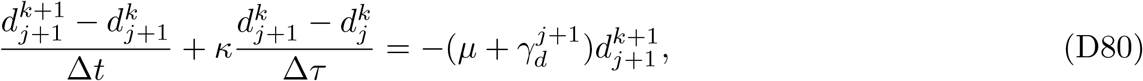

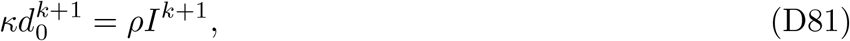

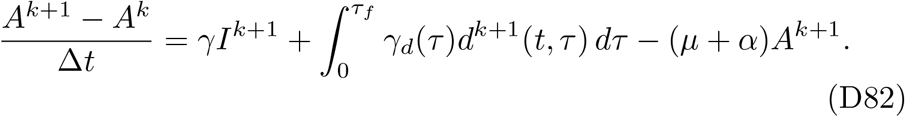

In this formulation, non-linear terms such as 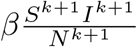 are approximated using known values from the previous iteration. This linearization (via Picard iteration) reduces computational complexity and allows for efficient solutions using standard linear solvers. To determine the discrete form of 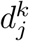, we first rewrite its governing equation.

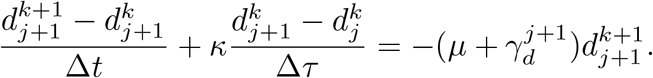

Using the substitution 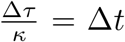, it follows that Δ*τ* = *κ*Δ*t*. Rewriting the equation, we obtain:

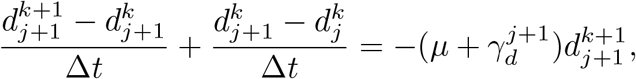

which simplifies to:

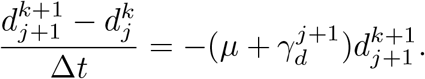

Rearranging the equations gives the following discrete update formulas for *S*^*k*+1^, *I*^*k*+1^, and *A*^*k*+1^.

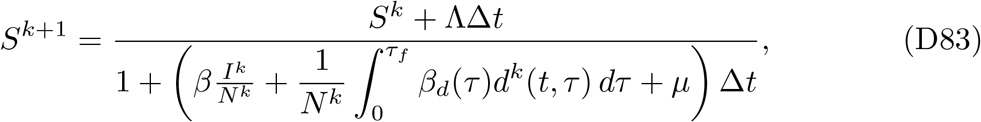

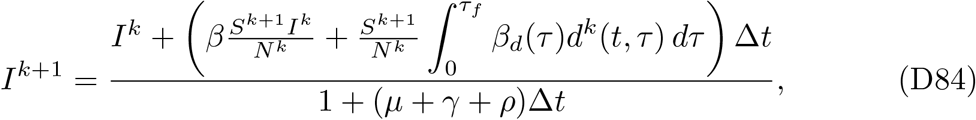

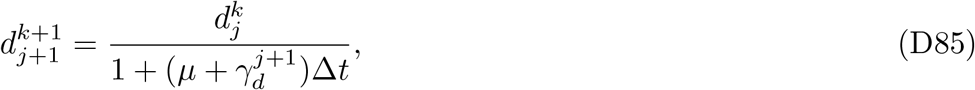

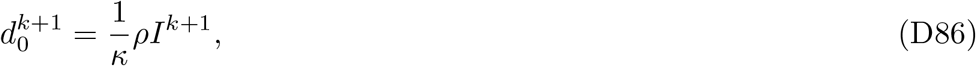

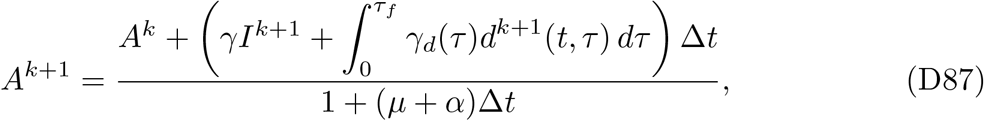

where 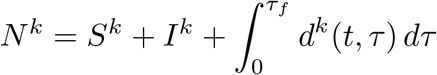

The initial conditions for the compartments 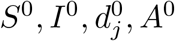 are set at the starting time *t*_0_, and are given by:

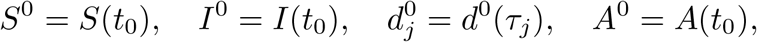

where *S*(*t*_0_), *I*(*t*_0_), *d*^0^(*τ*_*j*_), *A*(*t*_0_) represent the initial values of susceptible individuals, undiagnosed infected individuals, treated individuals in different treatment age groups, and individuals who have progressed to AIDS, respectively, at time *t*_0_. The integrals are approximated numerically using the trapezoidal rule in MATLAB.

## Appendix E Monte Carlo Simulations

The following are the steps for Monte Carlo Simulations.

- First, we solve the model equations numerically using parameter values obtained by fitting the model to the experimental data and record the predictions at the corresponding observation points.
- Next, we generate virtual datasets by adding measurement noise of varying magnitudes, *σ* ∈ *{*1%, 5%, 10%, 20%*}*, to the model predictions. The noise is assumed to follow a normal distribution with mean equal to the model output, 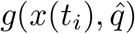, and standard deviation equal to *σ* times the model output, that is, 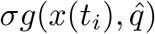.
- For each noise level, we create 1,000 synthetic datasets and reestimate the model parameters by minimizing the objective function for each dataset.
- The average relative estimation error (ARE) for each parameter is calculated as:

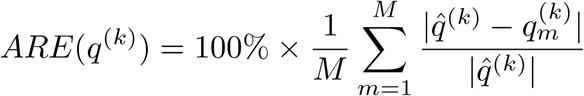

where 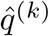 denotes the true value of the *k*^th^ parameter, and 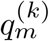 denotes the estimate of the *k*^th^ parameter obtained from the *m*^th^ Monte Carlo replicate.

## Appendix F Numerical Experiments

Figure F2 presents projections when both the transmission rate from diagnosed individuals, 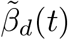, and the diagnosis rate, *ρ*(*t*), are refitted after 2019. This scenario provides the closest agreement with observed post-2019 trends, underscoring the combined importance of reducing transmission and expanding diagnosis coverage. HIV incidence continues to fall and meets the 2030 EHE incidence target, although the 2025 goal is not achieved, while the number of diagnoses decreases more gradually and does not meet the 2025 or 2030 diagnosis goals. The time-dependent parameters follow complementary trends: 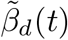 declines steadily after 2019, reflecting improved viral suppression, while *ρ*(*t*) increases, representing enhanced timely diagnosis. The combined effect produces a sustained reduction in the basic reproduction number *R*_0_(*t*) (Figure F2d). Numerically, *R*_0_ remains near 4.037 through 2019, declines to about 2.32 by 2025, and falls below the epidemic threshold by 2030 with *R*_0_ ≈ 0.898.

**Fig. F2.**
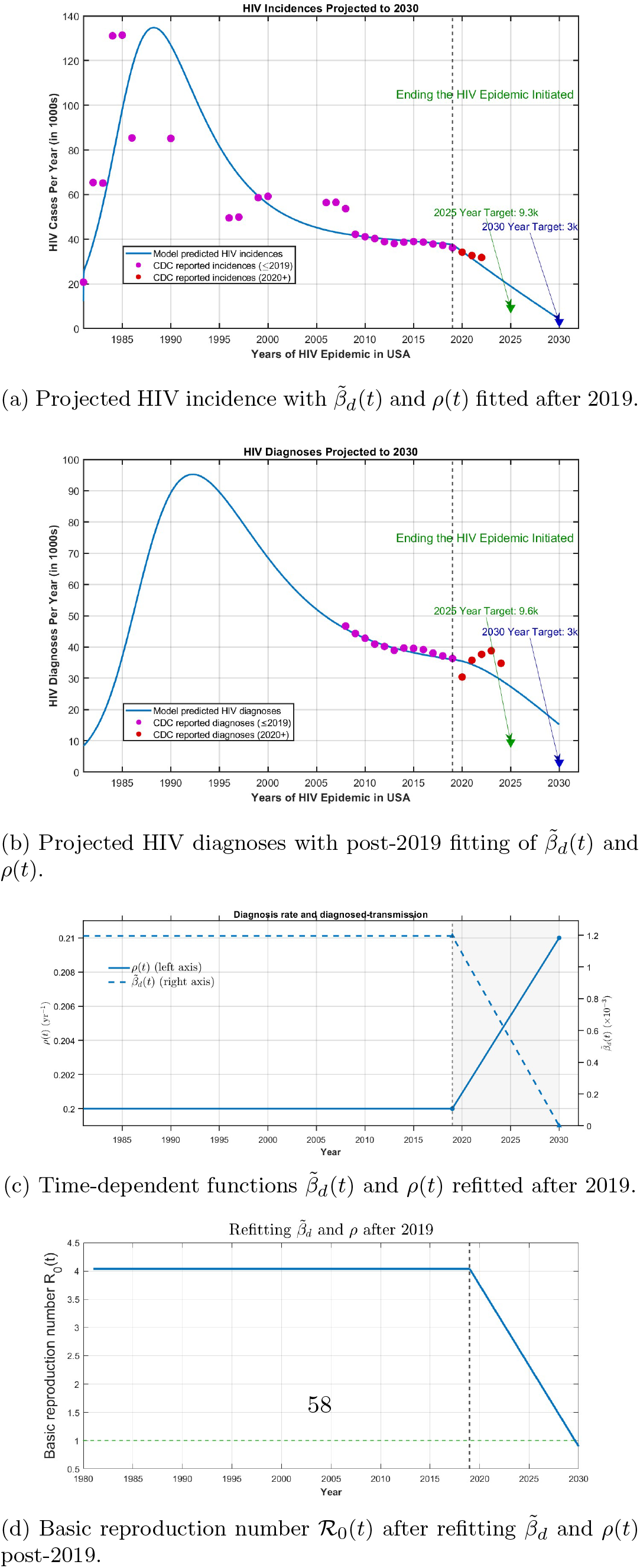
Model projections with both the diagnosed-transmission rate 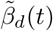 and the diagnosis rate *ρ*(*t*) refitted after 2019. Panels (a) and (b) show projected HIV incidence and diagnoses. Blue lines are model projections, purple dots are CDC data through 2019 used for fitting, and red dots are CDC data from 2020 onward used for validation. Th e vertical dashed line marks the 2019 launch of the Ending the HIV Epidemic initiative, and green arrows indicate the national targets for 2025 and 2030. panel (c) shows the refitted post-2019 functions 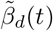 and *ρ*(*t*), and panel (d) shows the corresponding basic reproduction number *R*_0_(*t*).

## Notes

### Competing Interest Statement

The authors have declared no competing interest.

